# Genetic influences on alcohol flushing in East Asian populations

**DOI:** 10.1101/2023.04.28.23289268

**Authors:** Yoonsu Cho, Kuang Lin, Su-Hyun Lee, Canqing Yu, Dan Schmidt Valle, Daniel Avery, Jun Lv, Keumji Jung, Liming Li, George Davey Smith, China Kadoorie Biobank Collaborative Group, Zhengming Chen, Iona Y. Millwood, Gibran Hemani, Robin G. Walters

## Abstract

**Background:** Although it is known that variation in the *aldehyde dehydrogenase 2* (*ALDH2*) gene family influences the East Asian alcohol flushing response, knowledge about other genetic variants that affect flushing symptoms is limited.

**Methods:** We performed a genome-wide association study meta-analysis and heritability analysis of alcohol flushing in 15,105 males of East Asian ancestry (Koreans and Chinese) to identify genetic associations with alcohol flushing. We also evaluated whether self-reported flushing can be used as an instrumental variable for alcohol intake.

**Results:** We identified variants in the region of *ALDH2* strongly associated with alcohol flushing, replicating previous studies conducted in East Asian populations. Additionally, we identified variants in the alcohol dehydrogenase 1B (*ADH1B*) gene region associated with alcohol flushing. Several novel variants were identified after adjustment for the lead variants (*ALDH2*-rs671 and *ADH1B*-rs1229984), which need to be confirmed in larger studies. The estimated SNP-heritability on the liability scale was 13% (S.E. = 4%) for flushing, but the heritability estimate decreased to 6% (S.E. = 4%) when the effects of the lead variants were controlled for. Genetic instrumentation of higher alcohol intake using these variants recapitulated known associations of alcohol intake with hypertension. Using self-reported alcohol flushing as an instrument gave a similar association pattern of higher alcohol intake and cardiovascular disease-related traits (e.g. stroke).

**Conclusion:** This study confirms that *ALDH2*-rs671 and *ADH1B*-rs1229984 are associated with alcohol flushing in East Asian populations. Our findings also suggest that self-reported alcohol flushing can be used as an instrumental variable in future studies of alcohol consumption.

**Trial registration:** This study only used secondary data.

## Background

Alcohol flushing is a heritable condition in which a person develops flushes on the face or skin after drinking alcohol. Whilst pronounced alcohol flushing is rarely observed in Europeans, approximately 36% of East Asians experience alcohol flushing as well as other unpleasant symptoms (e.g. nausea and tachycardia) [1]. Previous genome-wide association studies (GWAS) identified two key genes associated with alcohol flushing, *alcohol dehydrogenase* 2 (*ALDH2*) and *aldehyde dehydrogenase* 1B (*ADH1B*) [2-4]. These genes encode enzymes that metabolize alcohol into acetaldehyde (*ADH1B*) and acetaldehyde into acetate (*ALDH2*). Genetic variants in *ALDH2* and *ADH1B* alter alcohol metabolism leading to prolonged, elevated levels of acetaldehyde. The excess acetaldehyde leads to physiological responses to alcohol consumption, including erythema on the face, nausea, and rapid heart rate [5, 6].

Most previous GWAS have focused on genetic associations with alcohol drinking status, rather than alcohol-induced responses, such as alcohol flushing [7, 8]. Candidate gene association studies have provided evidence for the association of *ALDH2* or *ADH1B* with alcohol flushing [9], but it is unclear whether there are loci other than *ALDH2* or *ADH1B* at which genetic variation appreciably influences flushing symptoms. Furthermore, investigations of putative causal genes for alcohol-related physiological responses have been conducted almost exclusively in individuals of European ancestry to date [7, 10], which risks missing variants with very low frequencies in European populations. Genetic biobanks from East Asian populations are growing in number, and with alcohol flushing highly prevalent amongst those participants there is an opportunity to improve our understanding of the relevant risk variants for the condition.

Recently, alcohol flushing has been proposed as a phenotypic instrumental variable (IV) for examining the health impacts of alcohol consumption in East Asian populations [11, 12]. Alcohol flushing is associated with lower levels of alcohol consumption and is assumed to be independent of confounders [13]. Considering the ease of including alcohol flushing questions in surveys compared with collecting genetic information, using flushing as an IV may be beneficial, enabling IV analysis in a simple, cost-effective, and non-invasive manner. Therefore, it would be helpful to fully understand the effects of genetic variants on alcohol flushing and to further characterise its utility as an IV.

In this study, we perform the largest GWAS of alcohol flushing to date, using 15,016 male individuals of East Asian ancestry from the China Kadoorie Biobank (CKB; N = 13,456) and the Korean Genome and Epidemiology Study (KoGES; N = 1,560). We also estimated the SNP-based heritability of alcohol flushing. Furthermore, we examined whether self-reported alcohol flushing can be used as a phenotypic IV for alcohol intake, comparing estimates with results from the genotypic IV (rs671 in *ALDH2*).

## METHODS

### Study population

This study was performed on two datasets, CKB (discovery set) and KoGES (replication set). CKB is a prospective study that recruited participants between 2004 and 2008. At baseline, 512,726 adults aged 30-79 years were recruited from 10 geographically defined regions of China (5 urban and 5 rural areas). All participants provide a 10mL blood sample which was processed into aliquots of buffy coat and plasma and stored at -70°C. Participants were prospectively followed up for cause-specific morbidity and mortality through linkage to death and disease registries and to the national health insurance system. Detailed information on the CKB is provided elsewhere [14, 15]. For the current analyses, we excluded individuals who were not genotyped or non-drinkers for whom information on alcohol flushing was not collected (Figure 1). Individuals with non-local ancestry were excluded from region-stratified GWAS analyses. Analyses were limited to male participants only since female participants’ alcohol intake is very low in China [16] and South Korea [17]. In total, 13,456 male CKB participants were included in regional GWAS analyses. For the meta-analysis, data for a total of 1,560 Korean men were obtained from KoGES [18]. For the IV analysis, we included 23,020 males from CKB who have information on alcohol flushing, alcohol intake amount and the known genetic instrument for alcohol (rs671 in *ALDH2*; Figure 1). All participants provided written informed consent approved by relevant local, national, and international ethics committees. Detailed information on the samples is provided in Supplementary Data.

**Figure 1.**
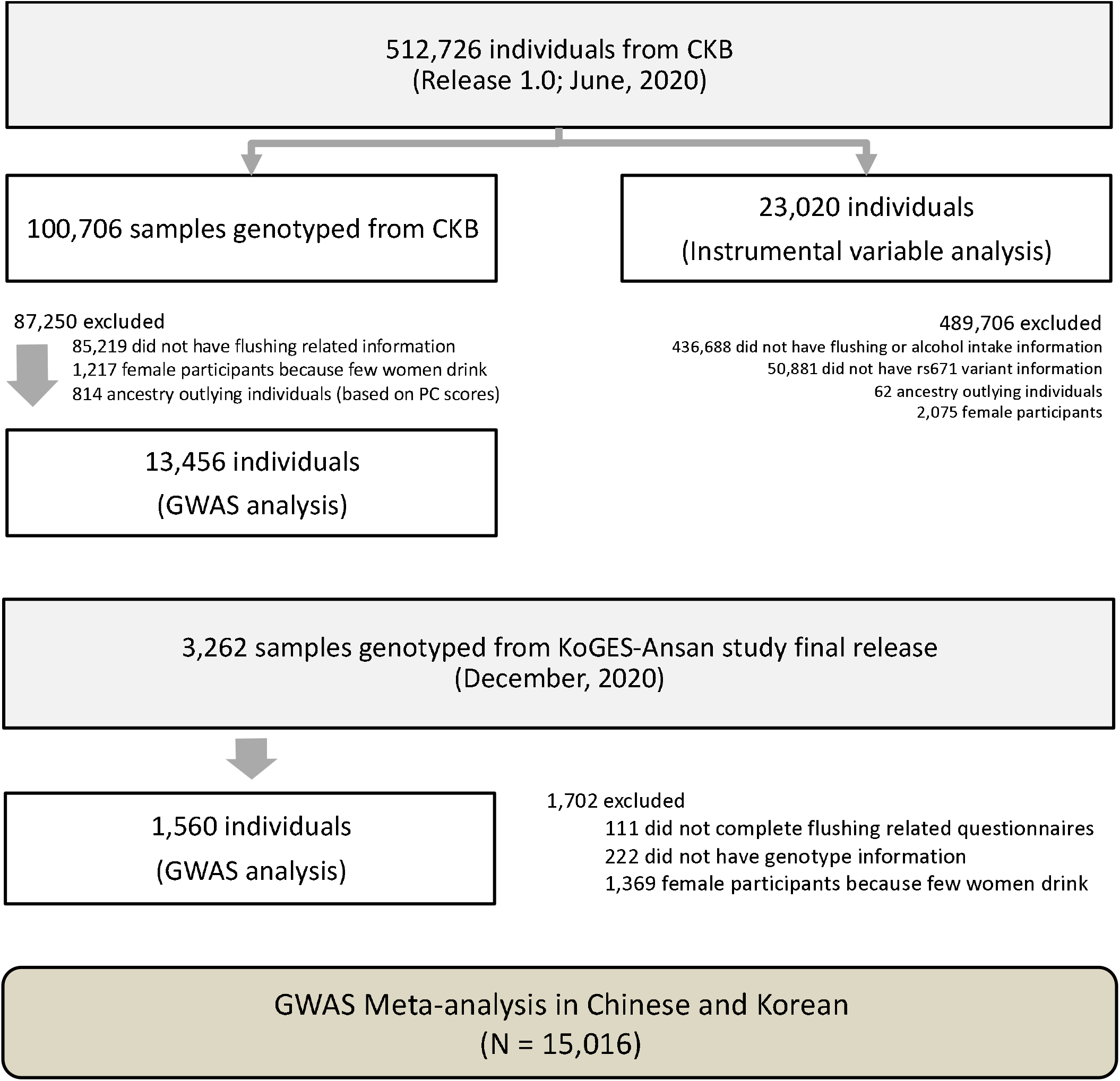
Flowchart of study population selection.

### Assessment of alcohol flushing and drinking patterns

In CKB, alcohol drinking patterns were investigated using interviewer-administered questionnaires. Participants were asked how often they had drunk alcohol during the previous 12 months (never or almost never; occasionally; only at certain seasons; every month but less than weekly; usually at least once a week). Based on the questionnaire, individuals who reported alcohol consumption in most weeks in the past year were identified as current drinkers. Current drinkers were asked further questions including types of beverage consumed, amount of alcohol drunk, and experience of flushing after drinking. Total alcohol intake (g/day) was calculated using the average alcohol content of each type of alcoholic beverage. Detailed information on the assessment of alcohol intake is available elsewhere [16, 19]. To investigate the presence of alcohol flushing symptoms among current drinkers, the following question was used: “Do you usually experience hot flushes or dizziness after drinking?” Participants were offered four options: “Yes, immediately”; “Yes, after a small amount of alcohol”; “Yes, but only after drinking a large amount of alcohol”, and “No”. Participants who experienced flushing immediately after drinking alcohol and those who flushed after a small amount of alcohol were classified as alcohol flushers. For sensitivity analyses, we defined alcohol flushing using different criteria (main, relaxed, strict, and continuous; see the Methods section in Supplementary Data for more details). All questionnaires were provided in Mandarin. The definition of flushing for KoGES is described in Supplementary Data.

### DNA sampling and genotyping

DNA was extracted from the buffy coat and was genotyped using the custom Affymetrix Axiom arrays and Illumina Golden Gate platform at BGI (Shenzhen, China), as previously described [15]. Data for a total of 100,706 individuals passed quality control criteria (call rate ≥ 95%, no sex mismatch, heterozygosity F statistic SD score <+3, no XY aneuploidy, no non-East Asian ancestry). Following variant QC (call rate > 0.98, no batch or plate effect, Hardy–Weinberg equilibrium P>10^−6^), imputation was performed using SHAPEITv3/IMPUTEv4 and the 1000 Genomes Project Phase 3 reference panel. After imputation, SNPs were removed if the MAF was low (< 0.01) or INFO was <0.3. After QC, 8,001,732 autosomal SNPs were used for association testing. Detailed information on the genotyping method and QC for KoGES is provided in Supplementary Data.

### Genome-wide association analyses

In CKB, genetic loci associated with flushing were investigated using BOLT-LMM v2.3.2 [20]. Three models were constructed. The first model was adjusted for age, age squared, the first ten genetic principal components (PCs), and genotyping array version (Model 1). We performed second and third GWAS analyses adjusting for the dosages of the SNPs that are known to be strongly associated with alcohol metabolism – rs671 in *ALDH2* (Model 2) and additionally rs1229984 in *ADH1B* (Model 3) [12]. We performed further GWAS analyses using different definitions of alcohol flushing (Supplementary Data). Each of the GWA analyses described above was performed separately for each geographical region (10 study areas). Within each region, SNPs with a low minor allele count (MAC < 6) or with Hardy– Weinberg equilibrium test values of P < 1 × 10^−6^ were excluded. Betas and standard errors (S.E.) obtained from BOLT-LMM were converted to log-odds ratios (OR) using log(OR) = β/(μ(1−μ)), where μ is the case-control ratio, following which region-level association statistics were combined using a fixed-effect inverse-variance-weighted meta-analysis using METAL [21]. One region (region 46, Liuzhou; n = 682) was excluded from the meta-analysis since the heritability estimate in this region was close to 0. We did not apply genomic control correction to the meta-analysis data because there was little evidence for inflation (all λ⍰<⍰1.008, Figure 2).

**Figure 2.**
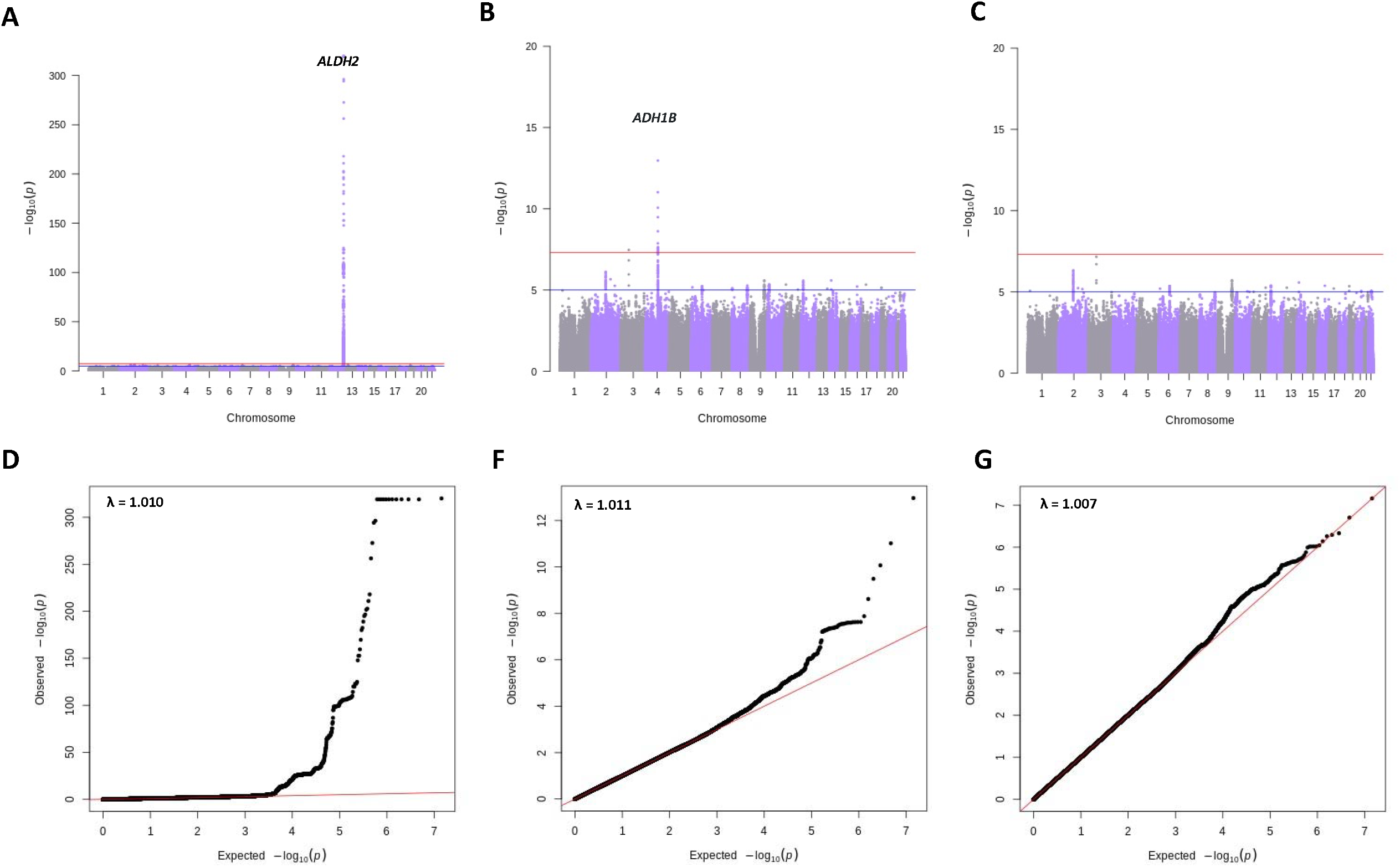
Manhattan plots and quantile-quantile for GWAS of flushing in Chinese population. Each plot represents the result from different models. (A) Model 1: controlling for age, age squared, PCs (1-10) (B) Model 2: covariates in model 1 plus *ALDH2* rs671 and (C) Model 3: covariates in model 2 plus *ADH1B* rs1229984. The y axis shows the age and sex adjusted -log10 P values, and the x axis presents positions along the chromosome (Chr.). The solid red line indicates the P value of 5 × 10^−8^ where the blue line indicates the P value of 1 × 10^−5^. (D-F) represent QQ plots for each model.

In KoGES, association tests were performed using PLINK 1.90 (available at https://www.cog-genomics.org/plink2). The GWA analysis of alcohol flushing was conducted using logistic regression assuming an additive genetic model using the three constructed models described above (Supplementary Data). SNPs with a low minor allele count (MAC < 20) were excluded.

For the GWAS meta-analysis of CKB and KoGES, We performed a fixed-effect inverse variance-weighted meta-analysis of the GWAS summary statistics from the CKB and KoGES using METAL [21].

The distributions of the observed P values of given SNPs were plotted against the theoretical distribution of expected P values to yield a quantile–quantile (QQ) plot for flushing (Figure 2). Regional association plots of the most significantly associated variants (P < 5 × 10^−8^) were generated by gassocplot2 (available at https://github.com/jrs95/gassocplot2) using the East Asian 1000-genome LD reference panel.

### Single nucleotide polymorphism-heritability analysis

The SNP heritability of alcohol flushing in the CKB sample was calculated using BOLT-REML, which provides a fast algorithm for multi-component modelling to partition SNP-heritability [22]. Heritability 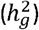 was estimated using the restricted maximum likelihood estimation method implemented in BOLT-REML. Since we defined alcohol flushing as a binary trait, we transformed the heritability on the observed scale to that on the liability scale 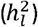 [23]. Analyses were adjusted for the covariates used in the GWAS analyses. SNP heritability in KOGES was estimated using the bivariate restricted maximum likelihood analysis implemented in GCTA [24, 25]. Detailed methods are described in the Supplementary Data.

### Mendelian randomisation analysis of alcohol flushing and disease outcomes

The causal effect of alcohol intake on blood pressure and cardiovascular diseases and related traits was evaluated using IV analyses with a two-stage least squares estimation method. A total of 23,020 individuals were included in the IV analyses (Figure 1). Self-reported alcohol flushing and the rs671 genotype were used as the phenotypic and genotypic instruments, respectively. We used the strict definition of flushing (i.e., immediately after consuming alcohol) as our IV. The magnitude of the association of alcohol intake (g/week) was scaled into a 280 g/week effect, as in a previous study [16]. For binary outcomes (i.e. stroke, myocardial infarction, coronary heart disease, hypertension, and diabetes), a two-stage logistic model was used. In the first stage, alcohol intake was instrumented by alcohol flushing or the rs671 genotype with adjustment for age, region, PCs (1-10), and genotyping array, using a linear regression model. In the second stage, the effect of alcohol on the risk of disease was estimated by fitting the alcohol intake value from the first stage, under a logistic regression model with adjustment for the same confounders as in the first stage. For continuous traits (i.e., aspartate aminotransferase [AST], gamma-glutamyl transferase [GTT], cholesterol, triglycerides, blood glucose, and blood pressure), a two-stage linear model was applied, similarly adjusting for confounders. Region-stratified analyses followed by meta-analysis gave similar results.

The values were reported as ORs per 280 g/week alcohol intake with 95% CIs for the binary outcomes and β-coefficients with 95% CIs for the continuous outcomes. We examined the strength and validity of each instrument using the F-statistic of the association of each instrument with alcohol intake (with an F-statistic >10 indicating adequate strength). The difference of estimates between instruments (alcohol flushing and rs671) was assessed using a difference of two means test [26].

## RESULTS

### General characteristics of the study population

The baseline characteristics of the study subjects according to flushing status are presented in Supplementary Table 1 and 2. In the CKB cohort, among 13,456 men with both alcohol flushing and genotype information, 17.9% reported flushing (i.e., flushing immediately after drinking alcohol or after drinking a small amount of alcohol). The mean weekly alcohol intake of non-flushers was 304.5 ± 259.0 g/week (mean ± standard deviation [SD]). Flushers had a lower mean weekly alcohol intake (228.1 ± 259.0 g/week) compared to non-flushers. Flushers had a higher proportion of rs671 A allele carriers (45.5 % vs 8.7 %) as well as rs1229984 A allele carriers (90.3 % vs 87.3 %) than non-flushers. The characteristics of 1,560 KoGES samples are described in Supplementary Table 2. Similar to the CKB, flushers in KoGES had a lower proportion of current drinkers who consumed a relatively small amount of alcohol compared to non-flushers.

### Genome-wide association analyses of flushing

In CKB, the top signal for GWAS of flushing (Model 1; See Methods) was at rs671, a functional variant in *ALDH2* (Beta = 2.86, S.E. = 0.07, P = 8.6 × 10^−416^; Figure 2 and Table 1; Supplementary Table 8). After adjustment for rs671 (Model 2), the strongest signal was detected at rs1229984 in *ADH1B* (Beta = 0.24, S.E = 0.03, P = 1.1 × 10^−13^; Supplementary Table 9). Additionally, model 2 identified a variant on chromosome 3 (rs1508403 in PTPRG, Beta = 0.84, S.E = 0.15, P = 3.38 × 10^−8^). There were no genome-wide significant SNPs after further adjustment for rs1229984 (Model 3; Figure 2).

**Table 1.** Top signals for the association with alcohol flushing in the CKB sample.

| Set | Model<sup>1</sup> | SNP<sup>2</sup> | CHR | Position (hg 19) | Nearest gene | A1 | A2 | EAF | Beta (S.E) | P-value<sup>3</sup> |
| --- | --- | --- | --- | --- | --- | --- | --- | --- | --- | --- |
| Main | Model 1 | rs671 | 12 | 112241766 | *ALDH2*<br>(missense) | A | G | 0.079 | 2.861 (0.066) | 8.61E-416 |
|  | Model 2 | rs1229984 | 4 | 100239319 | *ADH1B*<br>(missense) | T | C | 0.663 | 0.235 (0.032) | 1.08E-13 |
|  | Model 3 | rs1508403 | 3 | 62158555 | *PTPRG*<br>(intron) | T | C | 0.014 | 0.817 (0.152) | 6.92E-08 |
| Relaxed | Model 1 | rs671 | 12 | 112241766 | *ALDH2*<br>(missense) | A | G | 0.079 | 1.251 (0.050) | 1.91E-138 |
|  | Model 2 | rs1229984 | 4 | 100239319 | *ADH1B*<br>(missense) | T | C | 0.663 | 0.168 (0.026) | 4.71E-11 |
|  | Model 3 | rs148407052 | 7 | 76598533 | *LOC105375361*<br>(3KB Upstream) | A | G | 0.768 | 0.176 (0.035) | 5.117E-07 |
| Strict | Model 1 | rs671 | 12 | 112241766 | *ALDH2*<br>(missense) | A | G | 0.079 | 3.640 (0.105) | 4.21E-263 |
|  | Model 2 | rs1229984 | 4 | 100239319 | *ADH1B*<br>(missense) | T | C | 0.663 | 0.288 (0.052) | 2.56E-08 |
|  | Model 3 | rs150099059 | 1 | 211001286 | *KCNH1*<br>(Intron Variant) | C | G | 0.012 | 3.740 (0.651) | 9.40E-09 |
| Continuous | Model 1 | rs671 | 12 | 112241766 | *ALDH2*<br>(missense) | A | G | 0.079 | 0.925 (0.022) | 4.43E-380 |
|  | Model 2 | rs1229984 | 4 | 100239319 | *ADH1B*<br>(missense) | T | C | 0.663 | 0.090 (0.011) | 7.66E-17 |
|  | Model 3 | rs2903308 | 16 | 13656885 | *SHISA9*<br>(Non-coding) | G | A | 0.818 | 1.076 (0.205) | 1.44E-07 |

SNP, single nucleotide polymorphism; CHR, chromosome; A1/A2 alleles, minor and major alleles; EAF, effect allele frequency; S.E., standard error.
<sup>1</sup> Model 1: controlling for age, age squared, PCs (1-10); Model 2: covariates in model 1 plus *ALDH2* rs671; Model 3: covariates in model 2 plus *ADH1B* rs1229984.<sup>2</sup> Flushing and SNPs were regarded as dependent and independent variables, respectively. <sup>3</sup> The beta estimates, standard errors, and P-values were obtained from the linear regression.

**Table 2.**
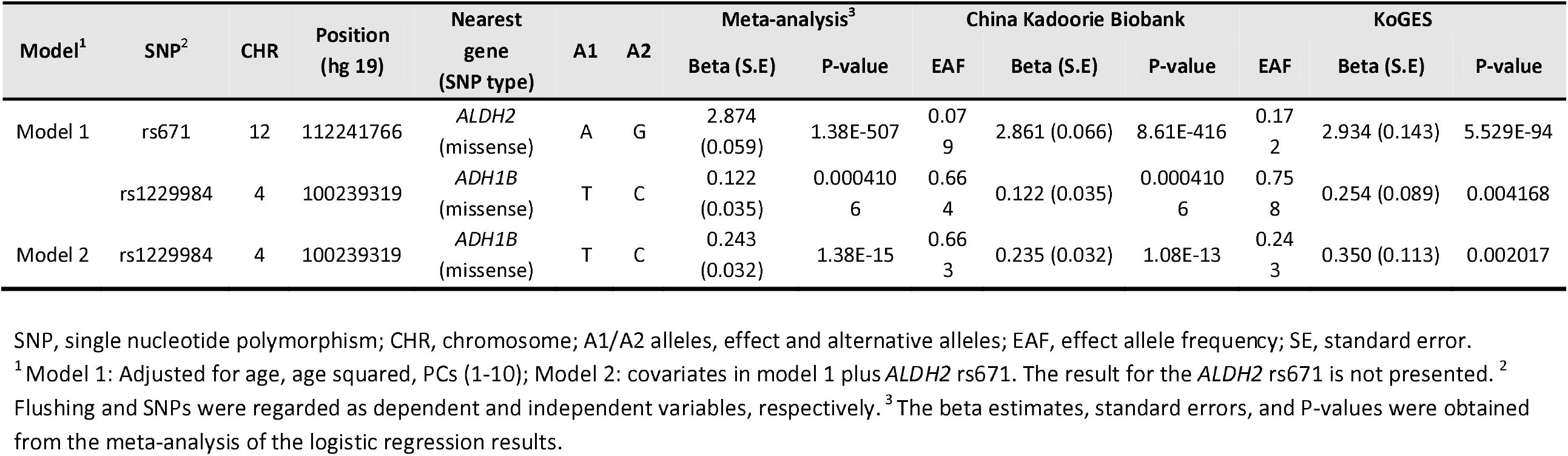
Genome-wide association results for 2 known alcohol-associated SNPs.

GWA analyses using different criteria for defining flushing showed no difference in the top signals for models 1 and 2 across the different definitions of flushing (see Supplementary Methods) although the P-values for the lead SNPs varied (Table 1; Supplementary Figure 1-3; Supplementary Table 10-16); The P values for the strongest signals became less significant for the relaxed flushing definition (ie., flushing after drinking any amount of alcohol) (Table 1; Supplementary Table 10-11). For the relaxed flushing definition, model 2 identified additional signals on chromosome 2 (rs532522882 *HPCAL1*; P = 1.29 × 10^−8^) along with the signal at *ADH1B* on chromosome 4 (Table 1; Supplementary Table 11). For the strict flushing definition (ie., flushing immediately after drinking alcohol), model 3 identified a few rare variants (MAF <= 0.01; Table 1 and Supplementary Table 14) that reached genome-wide significance including rs150099059 in KCNH1 (P = 9.4 × 10^−9^), rs1011755 on chromosome 11 (P = 1.6 × 10^−8^), and rs142761523 in CNTN (P = 2.6 × 10^−8^). For each flushing definition, model 3 also identified further suggestive associations marginally below the genome-wide significance threshold. These include rs148407052 in LOC105375361 (P = 5.1 × 10^−7^) for the relaxed flushing definition; and rs2903308 in SHISA9 (P = 1.4 × 10^−7^) for the continuous flushing definition. However, we were not able to replicate these findings in KoGES: either the association of these variants was not statistically significant, or they were not available in KoGES (Supplementary Table 6).

The GWAS results from an independent Korean cohort (KoGES) are presented in Supplementary Table 4. The GWAS identified strong association signals on chromosome 12 including rs671. In KoGES, *ADH1B* rs1229984 did not reach genome-wide significance across models 1-2. An apparent independent association at the chromosome 12 locus harbouring the *ALDH2* gene was identified after adjusting for rs671 (rs2074356, beta = 2.85, S.E = 0.26, 2.7 × 10^−28^; Supplementary Figure 4 and Supplementary Table 4), or adjusting for rs12231737, which was the top signal obtained from model 1 (rs2074356, beta = 2.26, S.E = 0.28, 2.9 × 10^−16^; Supplementary Table 4). To explore the obtained signals further, we conducted fine mapping using SuSiE which returned a single credible set. The credible set suggested that the conditionally independent signals are likely due to measurement error induced by relatively low imputation quality around the rs671 locus (data available on request).

### SNP heritability for alcohol flushing in the CKB and KoGES

SNP heritability of alcohol flushing among drinkers was estimated to be 12.6 % (SE = 4.0 %) on the liability scale 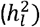 It decreased to 8.4 % (S.E. = 4.2 %) when we controlled for rs671 in *ALDH2* (Supplementary Table 5), and decreased further when we also controlled for rs1229984 in *ADH1B* 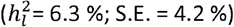, suggesting that rs671 and rs1229984 together explain half of the common variant genetic variance in alcohol flushing in Chinese males. SNP heritability estimates of alcohol flushing amongst drinkers and non-drinkers in the Korean population were imprecise due to the relatively small sample size but showed a pattern consistent with that seen in CKB.

### Using self-reported flushing as an instrumental variable

IV analyses among 23,020 men in CKB with flushing data showed that higher alcohol intake (as instrumented by absence of self-reported alcohol flushing) was nominally associated with a higher risk of intracerebral haemorrhage (OR per 280 g/week increase in alcohol intake = 3.28; 95% CI = 1.58 – 6.81), and total stroke (OR per 280 g/day increase in alcohol intake = 1.89; 95% CI = 1.28 – 6.81) as well as higher levels of AST, GGT, HDL cholesterol, log-transformed random blood glucose, and diastolic blood pressure (DBP; beta per 280 g/day increase in alcohol intake = 2.3 mm Hg; 95% CI = 0.9-3.7) (Table 3). These associations were generally consistent in direction and magnitude, although the estimates were more precise when using the rs671 genotype as an IV, which also provided evidence that higher alcohol intake caused a higher risk of hypertension and higher levels of systolic blood pressure (SBP), as well as increased risk of stroke types, coronary heart disease, and diabetes.

**Table 3.** Associations of alcohol intake (280 g/day) and diseases and traits using alcohol flushing or rs671 as instrumental variables in Chinese male drinkers.

| Instrument |  | Flushing<br>(N = 23,020) |  | ALDH2 rs671<br>(N = 23,020) |  | Heterogeneity<br>(P-value) |
| --- | --- | --- | --- | --- | --- | --- |
| Outcomes | N (case / control) | OR or beta coefficient (95% CI)<sup>1</sup> | P-value<sup>2</sup> | OR or beta coefficient (95% CI) | P-value<sup>2</sup> |  |
| Disease outcomes |  |  |  |  |  |  |
| Ischaemic stroke | 2,583 / 20,437 | 1.383 (0.882, 2.168) | 0.158 | 2.359 (1.616, 3.443) | 8.65e-06 | 0.957 |
| Intracerebral haemorrhage | 979 / 22,041 | 3.275 (1.575, 6.809) | 0.001 | 2.789 (1.535, 5.067) | 0.001 | 0.381 |
| Total stroke | 3,672 / 19,348 | 1.888 (1.277, 2.790) | 0.001 | 2.571 (1.857, 3.560) | 1.27e-08 | 0.880 |
| Myocardial infarction | 568 / 22,452 | 1.473 (0.620, 3.496) | 0.380 | 1.305 (0.648, 2.628) | 0.455 | 0.425 |
| Total coronary heart disease | 2,527 / 20,493 | 1.192 (0.767, 1.853) | 0.435 | 2.112 (1.455, 3.066) | 8.42e-05 | 0.968 |
| Hypertension | 2,727 / 20,293 | 1.489 (0.961, 2.307) | 0.075 | 2.135 (1.495, 3.049) | 3.03e-05 | 0.891 |
| Medical history |  |  |  |  |  |  |
| Self-reported hypertension | 8,920 / 14,100 | 1.019 (0.780, 1.330) | 0.892 | 2.089 (1.675, 2.605) | 6.17e-11 | 0.99995 |
| Self-reported diabetes | 732 / 22,288 | 1.494 (0.689, 3.239) | 0.310 | 4.663 (2.304, 9.438) | 1.86e-05 | 0.949 |
| Traits |  |  |  |  |  |  |
| Aspartate aminotransferase (μl/L) | 2,709 | 11.571 (0.182, 22.960) | 0.046 | 14.733 (6.472, 22.994) | 0.0005 | 0.670 |
| Gamma glutamyl transferase (μl/L) | 2,526 | 73.240 (6.853, 139.627) | 0.031 | 104.138 (55.010, 153.265) | 3.37e-05 | 0.768 |
| Cholesterol (mmol/L) | 2,710 | 0.207 (-0.208, 0.621) | 0.329 | 0.559 (0.244, 0.875) | 0.0005 | 0.907 |
| HDL cholesterol (mmol/L) | 2,710 | 0.172 (0.027, 0.316) | 0.020 | 0.113 (0.012, 0.215) | 0.0283 | 0.256 |
| LDL cholesterol (mmol/L) | 2,710 | -0.095 (-0.397, 0.207) | 0.538 | 0.101 (-0.117, 0.319) | 0.3655 | 0.849 |
| Triglyceride (mmol/L) | 2,710 | 0.382 (-0.528, 1.292) | 0.411 | 1.098 (0.416, 1.780) | 0.00162 | 0.891 |
| Log(Fasting blood glucose (mmol/L)) | 909 | -0.015 (-0.121, 0.091) | 0.780 | -0.007 (-0.083, 0.070) | 0.8594 | 0.548 |
| Log(Random blood glucose (mmol/L)) | 22,648 | 0.023 (0.008, 0.037) | 0.003 | 0.050 (0.038, 0.063) | 3.08e-15 | 0.997 |
| Systolic blood pressure (mmHg) | 23,020 | 1.090 (-1.358, 3.538) | 0.383 | 8.583 (6.536, 10.629) | 2.15e-16 | 0.999998 |
| Diastolic blood pressure (mmHg) | 23,020 | 2.295 (0.862, 3.728) | 0.002 | 5.857 (4.645, 7.068) | 3.01e-21 | 0.9999 |

<sup>1</sup> ORs and beta coefficients by instrumental variable (IV) estimation were obtained from IV regressions with a two-stage least squares estimation method (in logistic and linear regression models, respectively), using alcohol flushing as an instrument for alcohol intake. To predict the amount of alcohol intake (280 g/week), non-flushers were regarded as a reference group. <sup>2</sup> P values were derived from IV regression analysis with adjustment for age, region, and PC scores 1-10. The difference of estimates between instruments (alcohol flushing and rs671) was assessed using a difference of two means test.

## DISCUSSION

In this study, we investigated genetic variation associated with alcohol flushing and estimated the heritability of flushing in Chinese and Korean male populations. Strong signals were detected in *ALDH2* (Supplementary Figure 6) in both populations, supporting the previous evidence [27]. The SNP-based heritability estimate on the liability scale was 13% for flushing and decreased by 6% when the key variants (rs671 and rs1229984) were accounted for. The decrease in heritability supports the role of *ALDH2* and *ADH1B* as major contributors to the self-reported alcohol flushing response in the Chinese and Korean populations.

Among CKB male drinkers, 9% of non-flushers were *ALDH2*-rs671 A allele carriers compared with 46% of those reporting flushing. A similar pattern was observed in KoGES, where 9% of non-flushers were rs671-A carriers compared with 68% of flushers. To identify other variants that may influence alcohol flushing, we adjusted for the *ALDH2* rs671 genotype: this revealed a strong association of *ADH1B* rs1229984 with alcohol flushing. rs1229984 is a missense variant that has been extensively reported to be associated with alcohol consumption phenotypes such as alcohol intake status, and alcohol use disorders, including in European populations where the variant is present at low-frequency [28-30].

There has been some disagreement relating to the association of *ADH1B* with alcohol flushing. A low-dose alcohol challenge followed by a metabolite screen in Han Chinese men suggested that *ADH1B* did not associate with elevated blood acetaldehyde [31]. However, in a candidate gene study involving *ALDH2* and *ADH1B* in a sample of Japanese individuals with alcohol dependence, *ADH1B* did associate with flushing [32]. In CKB, the power to detect the *ADH1B* association is improved by reducing the residual variance after conditioning on rs671. However, the *ADH1B* association did not reach statistical significance in the Korean population. One theoretical explanation for that result is collider bias [33], in which flushing and *ADH1B* each influence alcohol dependence independently [32], and amongst cases become associated. Here, the *ADH1B* association is unlikely to arise due to this form of technical issue, because the association replicates in KoGES (albeit not at genome-wide significance) which has no alcohol consumption-related sample selection. Further GWAS in larger samples are required given the sample size of KoGES.

Several low-frequency variants were associated with different definitions of alcohol flushing in CKB (Table 1; Supplementary Tables 9-16), after controlling for the known variants (*ALDH2* rs671 and *ADH1B* rs1229984). These include PTPRG rs1508403 (MAF = 0.013) for the main flushing definition (Supplementary Table 9), *HPCAL1* rs532522882 (MAF = 0.004) and rs181957632 (MAF = 0.004) for the relaxed flushing definition (Supplementary Table 11), and KCNH1 rs150099059 (MAF = 0.01), and rs142761523 (MAF = 0.01) and rs144350123 in CNTN (MAF = 0.01) for the strict flushing definition (Supplementary Table 13). A GWAS study in 3,838 individuals of European- and African-American ancestry reported that the activities of PTPRG were associated with alcohol dependence [34]. A study in mice reported that the expression of *HPCAL1* was associated with alcohol consumption [35]. Furthermore, a study in rats reported that the KCNH1 gene, which encodes potassium voltage-gated channels, is differentially expressed in binge drinking groups [36]. The CNTN family has been suggested to be associated with alcohol independence by GWAS studies in European populations [37, 38]. Further studies with larger samples will be needed to replicate these findings.

SNP-based heritability analyses estimated that around 13% of the phenotypic variation in flushing is explained by common genetic variants. The heritability estimates decreased substantially when *ALDH2* rs671 was controlled for illustrating the strong effect of *ALDH2* on flushing in the Chinese population. These heritability estimates for flushing were much lower than all previous estimates for alcohol consumption [39]. One reason could be that our study only included regular drinkers. In this study, the subjects were asked about their experience of flushing based on their alcohol drinking status. This can be a source of selection bias where a sample can contain only those who report drinking. For example, individuals from CKB who do not regulary drink due to their knowledge of flushing are likely excluded from the current analysis. Also individuals who drink regardless of their flushing symptom may have developed compensatory feedback mechanisms [40], which can possibly contribute to weaker flushing symptoms. Consequently, this may lead to lower variance in flushing severity in the study subjects that could lead to lower heritability estimates in Chinese population.

The IV results demonstrated that self-reported alcohol flushing can be used successfully as an IV for alcohol consumption levels among drinkers. The pattern of associations of alcohol and disease traits was similar to a previous study in the Korean population that suggested the possibility of using self-reported alcohol flushing as an IV [11, 41]. However, we observed that the power to detect causal effects was generally attenuated in CKB when using self-reported flushing compared with the genetic IV, whereas the previous study by Cho et al. [41] demonstrated using self-reported alcohol flushing as an IV gave similar results to the use of the *ALDH2* rs671 variant as an IV. One major difference between the two studies is that CKB only had data available on alcohol flushing amongst individuals who self-reported regular drinking. Such structured sample selection can induce collider bias [33]. Indeed, in the CKB, the participants who regularly consumed alcohol had a lower prevalence of hypertension and lower BP levels than non-drinkers or ex-drinkers (Supplementary Table 7). This suggests that the IV analysis in CKB may have been affected by collider bias. For example, if higher levels of BP and flushing are both causally related to drinking, the association between alcohol intake and higher BP may be distorted (Supplementary Figure 7), given non-drinkers who flush were excluded from the current study. In this case, the genetic instrument (e.g. rs671) for the overall population is likely to be more reliable than a questionnaire as the genotypes are distributed completely randomly within the whole sample, regardless of their drinking status. By contrast, the self-reported IV based on the questionnaire is more likely to be subject to individuals’ drinking status.

This study has several other limitations. First, despite this being the largest genome-wide study of alcohol flushing to date, it is possible that there was limited statistical power to detect influential loci other than *ALDH2* and *ADH1B*. Second, our analyses included flushers who regularly drink, due to the design of the questionnaire used in CKB. Therefore, there is a possibility that those who do not drink alcohol due to their response to alcohol were not included in the current study. Nonetheless, results for our top loci are confirmed in two independent samples (Chinese and Koreans) showing that the identified genetic variants are likely to be strongly involved in flushing. Further GWAS and SNP heritability analyses are required in other East Asian populations. Third, some variants identified in CKB were relatively rare, and we could not test their association in KoGES, leaving the possibility that these variants were detected by chance. Fourth, although the variants used for GWAS were filtered to have high imputation scores (INFO >= 0.8), imputation accuracy using the 1000 genomes reference panel in Korean samples as was done for KoGES may still lead to measurement error. This is because, although the panel includes East Asian samples (Han Chinese and Japanese), it does not include Korean samples. It has been reported that the Korean population is genetically homogeneous due to geopolitical isolation, thus, Koreans genetically clustered distinctly from other East Asian populations [42]. Therefore, it could be speculated that while rs671 associated very strongly with flushing, it was not detected as the top signal at the *ALDH2* locus due to inaccuracy in imputation. Fifth, the use of alcohol flushing as an instrument may only reflect an effect of alcohol intake from a specific period of the life course (e.g. in adulthood) since alcohol flushing only occurs after an individual has started drinking (e.g. during adulthood).

## CONCLUSIONS

Despite these limitations, the results have epidemiologic and public health implications. Our findings underline the importance of additive genetic effects in modifying alcohol consumption behaviour and support the use of flushing or genetic variants (e.g. rs671 in *ALDH2*) as proxies for alcohol consumption in East Asian populations. To the best of our knowledge, this is the first GWAS to investigate putative causal variants for alcohol flushing and estimate the heritability of the condition in East-Asian populations.

## Data Availability

The datasets analysed during the current study are not publicly available due to institutional restrictions regarding accessibility, but are available from the corresponding author on reasonable request and with permission of the committee of CKB and KoGES.

## LIST OF ABBREVIATIONS

ALDH2: Aldehyde dehydrogenase 2
ADH1B: Alcohol dehydrogenase 1B
GWAS: Genome-wide association studies
IV: Instrumental variable
CKB: China Kadoorie Biobank
KoGES: Korean Genome and Epidemiology Study
PCs: Principal components
SE: Standard errors
OR: Odds ratios
QQ: quantile-quantile
SD: Standard deviation
AST: Aspartate aminotransferase
GGT: Gamma-glutamyl transferase
BP: Blood pressure
DBP: Diastolic blood pressure
SBP: Systolic blood pressure

## DECLARATIONS

### Ethics approval and consent to participate

Not applicable

### Consent for publication

Not applicable

### Author contributions

YC, IYM, GH, RGW and GDS conceptualized the project. YC, SHL, and KL performed statistical analyses. YC, IYM, GH, and RGW drafted the first version of the manuscript. All authors contributed to the interpretation of results and manuscript writing.

## Acknowledgements

China Kadoorie Biobank acknowledges the contribution of participants, project staff, the China National Centre for Disease Control and Prevention (CDC) and its regional offices.

## REFERENCES

1. Brooks PJ, Enoch MA, Goldman D, Li TK, Yokoyama A: The alcohol flushing response: an unrecognized risk factor for esophageal cancer from alcohol consumption. PLoS medicine 2009, 6(3):e50.

2. Edenberg HJ: The genetics of alcohol metabolism: role of alcohol dehydrogenase and aldehyde dehydrogenase variants. Alcohol research & health : the journal of the National Institute on Alcohol Abuse and Alcoholism 2007, 30(1):5–13.

3. Li D, Zhao H, Gelernter J: Strong association of the alcohol dehydrogenase 1B gene (ADH1B) with alcohol dependence and alcohol-induced medical diseases. Biological psychiatry 2011, 70(6):504–512.

4. Li D, Zhao H, Gelernter J: Strong protective effect of the aldehyde dehydrogenase gene (ALDH2) 504lys (*2) allele against alcoholism and alcohol-induced medical diseases in Asians. Human genetics 2012, 131(5):725–737.

5. Eng MY, Luczak SE, Wall TL: ALDH2, ADH1B, and ADH1C genotypes in Asians: a literature review. Alcohol research & health : the journal of the National Institute on Alcohol Abuse and Alcoholism 2007, 30(1):22–27.

6. Harada S, Agarwal DP, Goedde HW: Aldehyde dehydrogenase deficiency as cause of facial flushing reaction to alcohol in Japanese. Lancet 1981, 2(8253):982.

7. Clarke TK, Adams MJ, Davies G, Howard DM, Hall LS, Padmanabhan S, Murray AD, Smith BH, Campbell A, Hayward C et al: Genome-wide association study of alcohol consumption and genetic overlap with other health-related traits in UK Biobank (N=112 117). Mol Psychiatry 2017, 22(10):1376–1384.

8. Kranzler HR, Zhou H, Kember RL, Vickers Smith R, Justice AC, Damrauer S, Tsao PS, Klarin D, Baras A, Reid J et al: Genome-wide association study of alcohol consumption and use disorder in 274,424 individuals from multiple populations. Nature Communications 2019, 10(1):1499.

9. Macgregor S, Lind PA, Bucholz KK, Hansell NK, Madden PA, Richter MM, Montgomery GW, Martin NG, Heath AC, Whitfield JB: Associations of ADH and ALDH2 gene variation with self report alcohol reactions, consumption and dependence: an integrated analysis. Human molecular genetics 2009, 18(3):580–593.

10. Walters RK, Polimanti R, Johnson EC, McClintick JN, Adams MJ, Adkins AE, Aliev F, Bacanu SA, Batzler A, Bertelsen S et al: Transancestral GWAS of alcohol dependence reveals common genetic underpinnings with psychiatric disorders. Nat Neurosci 2018, 21(12):1656–1669.

11. Yun KE, Chang Y, Yun SC, Davey Smith G, Ryu S, Cho SI, Chung EC, Shin H, Khang YH: Alcohol and coronary artery calcification: an investigation using alcohol flushing as an instrumental variable. Int J Epidemiol 2017, 46(3):950–962.

12. Park BL, Kim JW, Cheong HS, Kim LH, Lee BC, Seo CH, Kang TC, Nam YW, Kim GB, Shin HD et al: Extended genetic effects of ADH cluster genes on the risk of alcohol dependence: from GWAS to replication. Hum Genet 2013, 132(6):657–668.

13. Zuccolo L, Holmes MV: Commentary: Mendelian randomization-inspired causal inference in the absence of genetic data. Int J Epidemiol 2016.

14. Chen Z, Chen J, Collins R, Guo Y, Peto R, Wu F, Li L, China Kadoorie Biobank collaborative g: China Kadoorie Biobank of 0.5 million people: survey methods, baseline characteristics and long-term follow-up. Int J Epidemiol 2011, 40(6):1652–1666.

15. Walters RG, Millwood IY, Lin K, Valle DS, McDonnell P, Hacker A, Avery D, Cai N, Kretzschmar WW, Ansari MA et al: Genotyping and population structure of the China Kadoorie Biobank. medRxiv 2022:2022.2005.2002.22274487.

16. Millwood IY, Walters RG, Mei XW, Guo Y, Yang L, Bian Z, Bennett DA, Chen Y, Dong C, Hu R et al: Conventional and genetic evidence on alcohol and vascular disease aetiology: a prospective study of 500 000 men and women in China. Lancet 2019, 393(10183):1831–1842.

17. Kang M, Min A, Min H: Gender Convergence in Alcohol Consumption Patterns: Findings from the Korea National Health and Nutrition Examination Survey 2007-2016. Int J Environ Res Public Health 2020, 17(24).

18. Kim Y, Han BG, Ko GESg: Cohort Profile: The Korean Genome and Epidemiology Study (KoGES) Consortium. Int J Epidemiol 2016.

19. Millwood IY, Li L, Smith M, Guo Y, Yang L, Bian Z, Lewington S, Whitlock G, Sherliker P, Collins R et al: Alcohol consumption in 0.5 million people from 10 diverse regions of China: prevalence, patterns and socio-demographic and health-related correlates. Int J Epidemiol 2013, 42(3):816–827.

20. Loh PR, Tucker G, Bulik-Sullivan BK, Vilhjalmsson BJ, Finucane HK, Salem RM, Chasman DI, Ridker PM, Neale BM, Berger B et al: Efficient Bayesian mixed-model analysis increases association power in large cohorts. Nat Genet 2015, 47(3):284–290.

21. Willer CJ, Li Y, Abecasis GR: METAL: fast and efficient meta-analysis of genomewide association scans. Bioinformatics 2010, 26(17):2190–2191.

22. Loh PR, Bhatia G, Gusev A, Finucane HK, Bulik-Sullivan BK, Pollack SJ, Schizophrenia Working Group of Psychiatric Genomics C, de Candia TR, Lee SH, Wray NR et al: Contrasting genetic architectures of schizophrenia and other complex diseases using fast variance-components analysis. Nat Genet 2015, 47(12):1385–1392.

23. Lee SH, Wray NR, Goddard ME, Visscher PM: Estimating missing heritability for disease from genome-wide association studies. Am J Hum Genet 2011, 88(3):294–305.

24. Yang J, Benyamin B, McEvoy BP, Gordon S, Henders AK, Nyholt DR, Madden PA, Heath AC, Martin NG, Montgomery GW et al: Common SNPs explain a large proportion of the heritability for human height. Nat Genet 2010, 42(7):565–569.

25. Lee SH, Yang J, Goddard ME, Visscher PM, Wray NR: Estimation of pleiotropy between complex diseases using single-nucleotide polymorphism-derived genomic relationships and restricted maximum likelihood. Bioinformatics 2012, 28(19):2540–2542.

26. Altman DG, Bland JM: Interaction revisited: the difference between two estimates. BMJ 2003, 326(7382):219.

27. Quillen EE, Chen XD, Almasy L, Yang F, He H, Li X, Wang XY, Liu TQ, Hao W, Deng HW et al: ALDH2 is associated to alcohol dependence and is the major genetic determinant of “daily maximum drinks” in a GWAS study of an isolated rural Chinese sample. Am J Med Genet B Neuropsychiatr Genet 2014, 165B(2):103–110.

28. Jorgenson E, Thai KK, Hoffmann TJ, Sakoda LC, Kvale MN, Banda Y, Schaefer C, Risch N, Mertens J, Weisner C et al: Genetic contributors to variation in alcohol consumption vary by race/ethnicity in a large multi-ethnic genome-wide association study. Mol Psychiatry 2017, 22(9):1359–1367.

29. Gelernter J, Kranzler HR, Sherva R, Almasy L, Koesterer R, Smith AH, Anton R, Preuss UW, Ridinger M, Rujescu D et al: Genome-wide association study of alcohol dependence:significant findings in African- and European-Americans including novel risk loci. Mol Psychiatry 2014, 19(1):41–49.

30. Liu MZ, Jiang Y, Wedow R, Li Y, Brazel DM, Chen F, Datta G, Davila-Velderrain J, McGuire D, Tian C et al: Association studies of up to 1.2 million individuals yield new insights into the genetic etiology of tobacco and alcohol use. Nature Genetics 2019, 51(2):237-+.

31. Peng GS, Chen YC, Wang MF, Lai CL, Yin SJ: ALDH2*2 but not ADH1B*2 is a causative variant gene allele for Asian alcohol flushing after a low-dose challenge: correlation of the pharmacokinetic and pharmacodynamic findings. Pharmacogenetics and genomics 2014, 24(12):607–617.

32. Yokoyama A, Yokoyama T, Kimura M, Matsushita S, Yokoyama M: Combinations of alcohol-induced flushing with genetic polymorphisms of alcohol and aldehyde dehydrogenases and the risk of alcohol dependence in Japanese men and women. PloS one 2021, 16(7):e0255276.

33. Munafo MR, Tilling K, Taylor AE, Evans DM, Davey Smith G: Collider scope: when selection bias can substantially influence observed associations. Int J Epidemiol 2018, 47(1):226–235.

34. Chen G, Zhang F, Xue W, Wu R, Xu H, Wang K, Zhu J: An association study revealed substantial effects of dominance, epistasis and substance dependence co-morbidity on alcohol dependence symptom count. Addict Biol 2017, 22(6):1475–1485.

35. Faccidomo S, Swaim KS, Saunders BL, Santanam TS, Taylor SM, Kim M, Reid GT, Eastman VR, Hodge CW: Mining the nucleus accumbens proteome for novel targets of alcohol self-administration in male C57BL/6J mice. Psychopharmacology (Berl) 2018, 235(6):1681–1696.

36. McClintick JN, McBride WJ, Bell RL, Ding ZM, Liu Y, Xuei X, Edenberg HJ: Gene Expression Changes in Glutamate and GABA-A Receptors, Neuropeptides, Ion Channels, and Cholesterol Synthesis in the Periaqueductal Gray Following Binge-Like Alcohol Drinking by Adolescent Alcohol-Preferring (P) Rats. Alcoholism, clinical and experimental research 2016, 40(5):955–968.

37. Clark SL, Aberg KA, Nerella S, Kumar G, McClay JL, Chen W, Xie LY, Harada A, Shabalin AA, Gao G et al: Combined Whole Methylome and Genomewide Association Study Implicates CNTN4 in Alcohol Use. Alcoholism, clinical and experimental research 2015, 39(8):1396–1405.

38. Oguro-Ando A, Zuko A, Kleijer KTE, Burbach JPH: A current view on contactin-4, -5, and -6: Implications in neurodevelopmental disorders. Mol Cell Neurosci 2017, 81:72–83.

39. Edwards AC, Deak JD, Gizer IR, Lai D, Chatzinakos C, Wilhelmsen KP, Lindsay J, Heron J, Hickman M, Webb BT et al: Meta-Analysis of Genetic Influences on Initial Alcohol Sensitivity. Alcoholism, clinical and experimental research 2018, 42(12):2349–2359.

40. Rutherford SL: From genotype to phenotype: buffering mechanisms and the storage of genetic information. Bioessays 2000, 22(12):1095–1105.

41. Cho Y, Kwak S, Lewis SJ, Wade KH, Relton CL, Smith GD, Shin MJ: Exploring the utility of alcohol flushing as an instrumental variable for alcohol intake in Koreans. Scientific reports 2018, 8(1):458.

42. Jeon S, Bhak Y, Choi Y, Jeon Y, Kim S, Jang J, Jang J, Blazyte A, Kim C, Kim Y et al: Korean Genome Project: 1094 Korean personal genomes with clinical information. Sci Adv 2020, 6(22):eaaz7835.

