## Supplementary Tables, Supplementary Figures for "Genetic influences on alcohol flushing in East Asian populations"

Yoonsu Cho et al.

### **Supplementary methods**

#### **China Kadoorie Biobank**

***Samples*** CKB is a prospective study that recruited between 2004 and 2008. At baseline, 512,891 adults aged 30-79 years were recruited from 10 geographically defined regions of China (5 urban and 5 rural areas). All participants are prospectively followed up for cause-specific mortality. Information on sociodemographic factors, medical history, smoking, drinking, diet, and physical activity were collected through interviewer-administered electronic questionnaire. Body weight, standing height, waist circumference, and blood pressure were measured by trained technicians while participants were wearing light clothes and no shoes. Fasting blood samples were collected and assayed for plasma HDL cholesterol, LDL cholesterol, triglycerides (TG), aspartate aminotransferase (AST), gamma glutamyl transferase (GGT), and blood glucose.

***Definition of Alcohol flushing for sensitivity GWAS analysis*** In the CKB, alcohol drinking patterns were investigated by self-reported questionnaire. The current drinkers were defined as individuals who drink more than once a week. The current drinkers were asked further questions: “do you usually experience hot flushes or dizziness after drinking?”. They were offered four options: “Yes, immediately”; “Yes, after small amount of alcohol”; “Yes, but only after drinking large amount of alcohol”, and “No”. Among the current drinkers, individuals who experience flushing immediately after drinking alcohol and those who flush after small amount of alcohol were classified as alcohol flushers (Main; Set 1). For sensitivity analysis, we used a range of criteria for alcohol flushing to consider alcohol sensitivity of the participants. First, we used a strict criterion for alcohol flushing classifying a group of participants who experience flushing immediately after drinking alcohol as flusher (Strict; Set 2). Second, we defined alcohol flusher using a relaxed criterion. A group of participants who experience flushing after drinking alcohol regardless of the amount alcohol was defined as flusher (Relaxed; Set 3). Flushing is also defined on a continuous scale ranging from 0 to 3, 0 corresponding to a "No" and 3 corresponding to an "immediately" (Continuous; Set 4). Independent variants were presented in the tables (Supplementary Table 8-19) after being clumped using the TwoSampleMR package (<https://mrcieu.github.io/TwoSampleMR/>) with an LD R^2^ threshold of 0.001 for SNPs within a 10,000 kilo base window and using the 1000 Genomes reference panel.

#### **Korean Genome and Epidemiology study (KoGES)**

***Samples*** Korean samples were obtained from the Ansan cohort study, which is part of the KoGES. Detailed information on the KoGES and Ansan cohort study are available elsewhere [1]. This study was based on the fourth set of follow-up data from the Ansan area alone because of the limited availability of data on alcohol flushing. Of 3,262 participants from the Ansan area, those who did not have data on alcohol flushing (n = 111), genotype (n = 222), drinking status (n = 0), or possible confounders (e.g., age, PCs; n = 0) were excluded. After exclusions, 1,560 unrelated men were included in the study. All participants signed an informed consent form that was approved by the Human Subjects Review Committees of Korea University Ansan Hospital (Ansan, South Korea) and Ajou University Medical Center (Suwon, South Korea). This study was approved by the Institutional Review Board of Yonsei University, Seoul, South Korea (Reference Number: 4-2015-1132). Demographic measurement such as including age, income, physical activity, and smoking status were collected through a health interview. Income level was divided into four groups (expressed in US currency): <1,000 USD (1,000,000 Korean Won); 1,000–2,000 USD; 2,000–4,000 USD; and ≥6,000 USD. Individuals who answered “Yes” to the following question were regarded as partaking in regular exercise: “Do you exercise at least once a week?” The subjects were asked to report how much time they spent during a typical day performing physical activities of five categories of intensity (sedentary, very light, light, moderate, and vigorous) and provide details on the activities corresponding to each category. Specific metabolic equivalent (MET) values were calculated by multiplying the time spent performing physical activities of each category to yield a total MET-hours score (0 for sleep or sedentary activity, 1.5 for very light activity, 2.5 for light activity, 5 for moderate activity, and 7.5 for vigorous activity). Subjects who smoked cigarettes during the survey period were defined as current smokers.

***Alcohol flushing and drinking patterns*** Based on the questionnaire, individuals who currently drank alcohol were categorized as current alcohol users. Ever drinkers were defined as individuals who had consumed alcohol at least once in their lifetimes. Total alcohol intake (g/day) was calculated using the average alcohol content of each type of alcoholic beverage. Detailed information on the calculation of alcohol content is available elsewhere [2]. Ever drinkers were asked further question: “Have you ever experienced facial flushing after consuming a little alcohol (e.g., a single cup [200 ml] of beer)?” Participants who answered “Yes” to the question were classified as flushers. All questionnaires were provided in Korean.

***DNA sampling and genotyping*** To genotype genomic DNA on the Genome-Wide Human SNP Array 5.0 (Affymetrix, Santa Clara, CA, USA), peripheral blood samples were collected from cohort participants at baseline. Detailed information on the genotyping method and quality control (QC) is provided elsewhere [3]. Briefly, genotype calls were made for 500,568 single nucleotide polymorphisms (SNPs) using Bayesian robust linear modeling with the Mahalanobis distance genotyping algorithm. After QC, markers with a high missing genotype call rate (> 5 %), low minor allele frequency (MAF; < 0.01), and significant deviation from the Hardy–Weinberg equilibrium (P < 1 × 10^−6^) were excluded, leaving 352,228 SNPs. The missing genotypes of individuals were imputed using IMPUTE software (available at https://mathgen.stats.ox.ac.uk/impute/impute_v2.html) based on the 1,000 genomes reference [4]. For post-imputation QC, SNPs were removed if the MAF was low (< 0.01), minor allele count was low (< 6), SNP call rate was low (<0.95), Hardy–Weinberg equilibrium was not satisfied (P < 1 × 10^−4^), or INFO was less than 0.8. After QC, a total of 6,461,358 autosomal SNPs were used for association testing.

***Genome wide association analysis and SNP-heritability analysis*** In KoGES, association tests were performed using PLINK 1.90 (available at <https://www.cog-genomics.org/plink2>). The GWA analysis of alcohol flushing was conducted using logistic regression with an additive genetic model with three constructed models. SNPs with a low minor allele count (MAC < 20) were excluded. The SNP heritability in Korean population was estimated using GCTA (available at cnsgenomics.com/software/gcta/) with bivariate restricted maximum likelihood analysis implemented in the software tool [5, 6]. A genetic relationship matrix (GRM) was calculated, and SNP liability was estimated using a linear mixed model, in which the GRM was included as a variance covariance matrix for a random effect. The model included the same covariates as in the three models described in the main method; Model 1 was adjusted for age, age squared, the first ten genetic principal components (PCs), and array information. Model 2 and 3 were adjusted for the covariates in the model 1 plus alcohol metabolism related genetic variants (the rs671 in *ALDH2* and the rs1229984 in *ADH1B*). As a sensitivity analysis, model 4 were additionally constructed for KoGES since the top signal in the model 1 was not rs671 in *ALDH2*. Model 4 was adjusted for the covariates in the model 1 plus the top signal obtained from model 1 (the rs12231737 in *TRAFD1*). To test whether the detected signals on chromosome 12 are independent from the rs671 in *ALDH2*, we conducted fine-mapping using SuSiE [7]. SuSiE obtains credible sets of variants with 95% cumulative posterior probability through Iterative Bayesian Stepwise Selection. 1000-genome LD reference panel for East Asian population was used for SuSiE.

### **Supplementary Results**

#### **Genome-wide association analyses of flushing in KoGES**

The GWA results from an independent Korean cohort are presented in Supplementary Table 5. The GWAS identified strong association signals on chromosome 12. The strongest association was detected for rs12231737 in TRAFD1 (Beta = 3.28, S.E. = 0.15, P = 2.7 x 10^-100^). After adjustment for the rs671 variant, another strong signal was observed for rs20074356 in HECTD4 (Beta = 2.84, S.E. = 0.03, P = 5.4 x 10^-28^), which is in LD with ALDH2. This signal remained after further adjustment for the rs1229984 variant in ADH1B (P = 3.7 x 10^-28^). In KoGES, ADH1B rs1229984 did not reach genome wide significance across the model 1-3.

**Meta-analysis of results for the known alcohol variants from the CKB and KoGES**

Our GWAS meta-analysis confirmed the alcohol metabolism-associated loci previously identified among individuals of East Asian ancestry (Table 2; Supplementary Figure 5). The strongest association signal from the meta-analysis of the two studies was observed for rs671 in *ALDH2* (Beta = 2.87, S.E. = 0.07, P = 1.4 x 10^-507^). The rs1229984 variant in ADH1B, a missense variant known to be involved in alcohol metabolism, reached genome-wide significance after conditioning on rs671 (Beta = 0.24, S.E. = 0.03, P = 1.4 x 10^-15^).

A summary of the strongest association signals from each model are presented in Supplementary Table 3. For model 1, genome-wide significant associations (p < 5 x 10^-8^) were obtained for 1,768 SNPs mapped to chromosome 12 (Figure 2; Supplementary Table 17-19). Note that the strong signal on chromosome 12 in model 3 was derived exclusively from KoGES, and is potentially an artefact derived from poor imputation of rs671, as noted above.

### **Supplementary Table**

#### **Supplementary Table 1. Characteristics of the study subjects according to alcohol flushing status in CKB.**

|  | **Current drinkers** | | | | |
| --- | --- | --- | --- | --- | --- |
|  | **Total**  **(n=13,456)** |  | **Men** | | |
| **Variables** |  |  | **Non-alcohol flushers**  **(n= 11,054)** |  | **Alcohol**  **flushers**  **(n= 2,402)** |
| Age (years ± S.D.) | 53.0 ± 10.6 |  | 53.1 ± 10.7 |  | 52.7 ± 10.6 |
| Region (%) |  |  |  |  |  |
| Urban | 53.4 |  | 51.9 |  | 57.7 |
| Rural | 46.6 |  | 48.1 |  | 42.3 |
| Monthly household income (yuan, %) |  |  |  |  |  |
| <10,000 | 24.2 |  | 23.2 |  | 28.4 |
| 10,000-19,999 | 29.0 |  | 29.4 |  | 26.9 |
| 20,000–34,999 | 25.8 |  | 26.2 |  | 23.9 |
| ≥35,000 | 21.1 |  | 21.2 |  | 20.9 |
| Drinking |  |  |  |  |  |
| Total alcohol intake (g/week ± S.D.) | 290.9 ± 250.8 |  | 304.5 ± 259.0 |  | 228.1 ± 196.6 |
| Smoking (%) |  |  |  |  |  |
| Non-smoker | 7.8 |  | 7.7 |  | 8.3 |
| Ex-smoker | 13.3 |  | 13.0 |  | 14.7 |
| Current smoker | 78.8 |  | 79.2 |  | 77.0 |
| Physical activity |  |  |  |  |  |
| MET-hours (hour/day ± S.D.) | 6.2 ± 3.5 |  | 6.2 ± 3.5 |  | 6.2 ± 3.4 |
| Adult height (cm ± S.D.) | 165.5 ± 6.8 |  | 165.7 ± 6.8 |  | 164.5 ± 7.0 |
| Adult weight (kg ± S.D.) | 65.0 ± 11.6 |  | 65.1 ± 11.6 |  | 64.5 ± 11.3 |
| Genotype (%) |  |  |  |  |  |
| ADH1B rs1229984 (AA/AG/GG) | 45.2 / 42.6 / 12.2 |  | 44.8 / 42.5 / 12.7 |  | 47.1 / 43.3 / 9.7 |
| ALDH2 rs671(GG/GA+AA) | 84.8 / 15.2 |  | 91.4 / 8.7 |  | 54.6 / 45.5 |

S.D., standard deviation; MET, metabolic equivalent.

Values are presented as mean ± standard deviation for continuous variables, or number (percentages) for categorical variables. P values are not presented as the aim of this analysis was to show the basic characteristics of the study subjects rather than to perform significance testing.

#### **Supplementary Table 2. Characteristics of study subjects according to alcohol flushing status in KoGES.**

|  | **Ever drinkers** | | | |
| --- | --- | --- | --- | --- |
|  | **Total**  **(n=1,560)** |  | **Men** | |
| **Variables** |  |  | **Non-alcohol flushers**  **(n=946)** | **Alcohol flushers**  **(n=614)** |
| Age (years ± S.D.) | 56.4 ± 7.2 |  | 55.8 ± 6.9 | 57.3 ± 7.6 |
| Monthly household income (won, %) |  |  |  |  |
| <1,000,000 | 9.1 |  | 8.3 | 10.3 |
| 1,000,000-2,000,000 | 14.2 |  | 11.6 | 18.1 |
| 2,000,000-4,000,000 | 41.0 |  | 41.9 | 39.9 |
| ≥4,000,000 | 35.8 |  | 38.5 | 31.7 |
| Drinking |  |  |  |  |
| Current drinker (%) | 74.9 |  | 86.7 | 56.8 |
| Total alcohol intake (g/day ± S.D.) | 23.3 ± 27.0 |  | 26.7 ± 27.9 | 14.9 ± 22.7 |
| Smoking (%) |  |  |  |  |
| Non-smoker | 21.2 |  | 20.1 | 23.0 |
| Ex-smoker | 51.1 |  | 50.7 | 51.6 |
| Current smoker | 27.7 |  | 29.2 | 25.4 |
| Physical activity |  |  |  |  |
| MET hours (hour/day ± S.D.) | 6.72 ± 1.50 |  | 6.67 ± 1.50 | 6.79 ± 1.51 |
| Adult height (cm) | 167.4 ± 5.6 |  | 167.6 ± 5.5 | 167.1 ± 5.6 |
| Adult weight (kg ± S.D.) | 69.2 ± 9.0 |  | 69.7 ± 9.1 | 68.4 ± 8.9 |
| Genotype (%) |  |  |  |  |
| ADH1B rs1229984 (TT/TC/CC) | 58.1 / 36.0 / 5.9 |  | 55.6 / 37.6 / 6.8 | 62.1 / 33.4 / 4.6 |
| ALDH2 rs671(GG/GA/AA) | 67.6 / 29.6 / 2.8 |  | 90.9 / 8.7 / 0.4 | 31.6 / 61.9 / 6.5 |

S.D., standard Deviation; MET, metabolic equivalent.

Values are presented as mean ± standard deviation for continuous variables, or number (percentages) for categorical variables. P values are not presented as the aim of this analysis was to show the basic characteristics of the study subjects rather than to perform significance testing.

#### **Supplementary Table 3. Most significant SNPs from meta-analysis.**

| **Top loci identified in the meta-analysis** | | | | | | | | | | | | | | |
| --- | --- | --- | --- | --- | --- | --- | --- | --- | --- | --- | --- | --- | --- | --- |
| **Model^1^** | **SNP** | **CHR** | **Position**  **(hg 19)** | **Nearest gene**  **(SNP type)** | **A1** | **A2** | **Meta-analysis** | | **China Kadoorie Biobank** | | | **KoGES** | | |
|  |  |  |  |  |  |  | **Beta (S.E)^2^** | **P-value^2^** | **EAF** | **Beta (S.E)^2^** | **P-value^2^** | **EAF** | **Beta (S.E)^2^** | **P-value^2^** |
| Model 1 | rs671 | 12 | 112241766 | ALDH2  (missense) | A | G | 2.874 (0.059) | 1.38E-507 | 0.079 | 2.861 (0.066) | 8.61E-416 | 0.172 | 2.934 (0.143) | 5.529E-94 |
| Model 2 | rs1229984 | 4 | 100239319 | ADH1B  (missense) | T | C | 0.243 (0.032) | 1.38E-15 | 0.663 | 0.235 (0.032) | 1.08E-13 | 0.243 | 0.350 (0.113) | 0.002017 |
| Model 3 | rs11066132 | 12 | 112428206 | NAA25  (intron) | T | C | 1.673 (0.209) | 1.294E-15 | 0.080 | -0.144 (0.233) | 0.5359 | 0.169 | 2.763 (0.298) | 2.092E-20 |

SNP, single nucleotide polymorphism; CHR, chromosome; A1/A2 alleles, minor and major alleles; EAF, effect allele frequency; SE, standard error.

^1^ Model 1: controlling for age, age squared, PCs (1-10); Model 2: covariates in model 1 plus ALDH2 rs671; Model 3: covariates in model 2 plus ADH1B rs1229984. ^2^ The beta estimates, standard errors, and P-values were obtained from the meta-analysis. ^3^ KCNIP4 rs189252575 did not exist in KoGES.

#### **Supplementary Table 4. Top single nucleotide polymorphism (SNP) that presented the smallest adjusted P values in additive-model analyses for the association with alcohol flushing in KoGES.**

| **Model^1^** | **SNP^2^** | **CHR** | **Position (hg 19)** | **Nearest gene** | **A1** | **A2** | **EAF** | **Beta (S.E)** | **P-value^3^** |
| --- | --- | --- | --- | --- | --- | --- | --- | --- | --- |
| Model 1 | rs12231737 | 12 | 112574616 | *TRAFD1*  (intron) | T | C | 0.176 | 3.283 (0.154) | 2.749E-100 |
| Model 2 | rs2074356 | 12 | 112645401 | *HECTD4*  (intron) | A | G | 0.149 | 2.849 (0.258) | 2.746E-28 |
| Model 3 | rs2074356 | 12 | 112645401 | *HECTD4*  (intron) | A | G | 0.154 | 2.878 (0.261) | 2.478E-28 |
| Model 4 | rs2074356 | 12 | 112645401 | *HECTD4*  (intron) | A | G | 0.154 | 2.260 (0.276) | 2.885E-16 |

SNP, single nucleotide polymorphism; CHR, chromosome; A1/A2 alleles, minor and major alleles; EAF, effect allele frequency; S.E., standard error. ^1^ Model 1: controlling for age, age squared, PCs (1-10); Model 2: covariates in model 1 plus *ALDH2* rs671; Model 3: covariates in model 2 plus *ADH1B* rs1229984; Model 4: covariates in model 1 plus *TRAFD1* rs12231737.^2^ Flushing and SNPs were regarded as dependent and independent variables, respectively. ^3^ The beta estimates, standard errors, and P-values were obtained from the logistic regression results.

#### **Supplementary Table 5. Estimates of total SNP heritability.**

|  | $\boldsymbol{h}_{\boldsymbol{l}}^{\boldsymbol{2}}$ **% (S.E.)** | |
| --- | --- | --- |
| **Model^1^** | **CKB** | **KoGES** |
| Model 1 | 12.6 (4.0) | 111.8 (30.1) |
| Model 2 | 8.4 (4.2) | 43.6 (31.0) |
| Model 3 | 6.3 (4.2) | 42.3 (31.1) |

S.E., standard error. ^1^ Model 1: controlling for age, age squared, PCs (1-10); Model 2: covariates in model 1 plus ALDH2 rs671; Model 3: covariates in model 2 plus ADH1B rs1229984. ^2^ Heritability ($h_{l}^{2}$) was estimated using restricted maximum likelihood estimation method implemented in BOLT-REML for CKB and GCTA for KoGES, respectively. Obtained SNP heritability values were transformed to the liability scale.

#### **Supplementary Table 6. Replication of the novel variants from CKB in KoGES.**

| **Set** | **Model^1^** | **SNP^2^** | **CHR** | **Position (hg 19)** | **Nearest gene** | **A1** | **A2** | **EAF** | **Beta (S.E)** | **P-value^3^** |
| --- | --- | --- | --- | --- | --- | --- | --- | --- | --- | --- |
| **Main** | Model 2 | rs1508403 | 3 | 62158555 | *PTPRG*  (intron) |  |  |  | Not available |  |
|  | Model 3 | rs1508403 | 3 | 62158555 | *PTPRG*  (intron) |  |  |  | Not available |  |
| **Relaxed** | Model 2 | rs532522882 | 2 | 10486609 | *HPCAL1*  (Intron Variant) |  |  |  | Not available |  |
|  |  | rs181957632 | 2 | 10483567 | *HPCAL1*  (Intron Variant) |  |  |  | Not available |  |
|  | Model 3 | rs148407052 | 7 | 76598533 | *LOC105375361*  (3KB Upstream) |  |  |  | Not available |  |
| **Strict** | Model 3 | rs150099059 | 1 | 211001286 | *KCNH1*  (Intron Variant) | C | G | 0.0105 | -0.584 (0.514) | 0.2554 |
|  |  | rs149732562 | 1 | 211050104 | *KCNH1*  (Intron Variant) | A | G | 0.0101 | -0.584 (0.514) | 0.255 |
|  |  | rs1011755 | 11 | 80185799 | - | A | C | 0.014 | 0.041 (0.427) | 0.923 |
|  |  | rs1011756 | 11 | 80185503 | - | A | T | 0.014 | 0.041 (0.427) | 0.923 |
|  |  | rs142761523 | 3 | 1259893 | *CNTN*  (Intron Variant) |  |  |  | Not available |  |
|  |  | rs144350123 | 3 | 1270101 | *CNTN*  (Intron Variant) |  |  |  | Not available |  |
|  |  | rs189057628 | 3 | 1290460 | *CNTN*  (Intron Variant) |  |  |  | Not available |  |
| **Continuous** | Model 3 | rs2903308 | 16 | 13656885 | *SHISA9*  (Non-coding) | A | G | 0.191 | 0.065 (0.118) | 0.5837 |

SNP, single nucleotide polymorphism; CHR, chromosome; A1/A2 alleles, minor and major alleles; EAF, effect allele frequency; S.E., standard error.

^1^ Model 1: controlling for age, age squared, PCs (1-10); Model 2: covariates in model 1 plus *ALDH2* rs671; Model 3: covariates in model 2 plus *ADH1B* rs1229984.^2^ Flushing and SNPs were regarded as dependent and independent variables, respectively. ^3^ The beta estimates, standard errors, and P-values were obtained from the linear regression.

#### **Supplementary Table 7. Comparison of basic characteristics between non-male drinkers and male drinkers in CKB.**

|  | **Study subjects** | | | | | |
| --- | --- | --- | --- | --- | --- | --- |
|  | **Never drinkers**  **(n = 42,779)** |  | **Ex-drinkers**  **(n = 7,924)** |  | **Current drinkers**  **(n = 159,501)** | **P value^1^** |
| Age (years ± S.D.) | 56.1 ± 11.1 |  | 59.4 ± 9.9 |  | 51.6 ± 10.6 | <2e-16 |
| Genotype (%) |  |  |  |  |  |  |
| ADH1B rs1229984 (AA/AG/GG) | 50.1/42.7/7.2 |  | 49.9/40.4/9.7 |  | 47.0/42.9/10.1 | < 1E-27 |
| ALDH2 rs671(GG/GA/AA) | 31.1/52.5/16.4 |  | 72.3/26.8/0.9 |  | 71.5/27.1/1.4 | 2.38E-27 |
| BMI (kg/m^2^) |  |  |  |  |  |  |
| Disease (incidence, %) |  |  |  |  |  |  |
| Ischaemic stroke | 15.5 |  | 16.2 |  | 10.0 | 1.73E-90 |
| Intracerebral haemorrhage | 17.6 |  | 5.1 |  | 2.0 | 1.73E-90 |
| Total stroke | 14.4 |  | 22.3 |  | 12.4 | 1.73E-90 |
| Myocardial infarction | 17.8 |  | 3.98 |  | 1.82 | 1.73E-90 |
| Total coronary heart disease | 15.2 |  | 18.0 |  | 9.74 | 1.73E-90 |
| Hypertension | 14.5 |  | 21.8 |  | 9.68 | 1.73E-90 |
| Disease (prevalence, %) |  |  |  |  |  |  |
| Self-reported hypertension | 10.5 |  | 43.1 |  | 32.3 | 1.73E-90 |
| Self-reported diabetes | 17.6 |  | 5.21 |  | 3.06 | 1.73E-90 |
| Traits (years ± S.D.) |  |  |  |  |  |  |
| Aspartate aminotransferase (µl/L) | 30 ± 15.1 |  | 29 ± 14.4 |  | 30.1 ± 23.7 | 0.776 |
| Gamma glutamyl transferase (µl/L) | 27.7 ± 32.4 |  | 33.3 ± 70.5 |  | 47 ± 107.9 | 6.97E-17 |
| Cholesterol (mmol/L) | 4.4 ± 0.9 |  | 4.5 ± 0.9 |  | 4.6 ± 1 | 1.16E-20 |
| HDL cholesterol (mmol/L) | 1.2 ± 0.3 |  | 1.2 ± 0.3 |  | 1.2 ± 0.3 | 1.02E-12 |
| LDL cholesterol (mmol/L) | 2.3 ± 0.7 |  | 2.4 ± 0.7 |  | 2.3 ± 0.7 | 0.0000004 |
| Triglyceride (mmol/L) | 1.7 ± 1.3 |  | 1.8 ± 1.4 |  | 2.1 ± 1.9 | 2.08E-14 |
| Fasting blood glucose (mmol/L) | 6.8 ± 2.9 |  | 7 ± 2.6 |  | 7 ± 2.5 | 0.0626 |
| Random blood glucose (mmol/L) | 6 ± 2.5 |  | 6.4 ± 2.9 |  | 5.9 ± 2.3 | 1.97E-21 |
| Systolic blood pressure (mmHg) | 133.8 ± 21.5 |  | 137.9 ± 22.8 |  | 132.3 ± 19.4 | 1.76E-61 |
| Diastolic blood pressure (mmHg) | 78.3 ± 11.5 |  | 79.9 ± 12.2 |  | 79.4 ± 11.3 | 2.73E-70 |

^1^ Values are presented as mean ± standard deviation for continuous variables, or number (percentages) for categorical variables. ^1^P values were derived from One way ANOVA test (continuous variables) or Chi-square test (categorical variables) for differences between flushers and non-flushers. Statistical significance was defined as p <0.05.

### **Figure legends**

**Supplementary Figure 1. Manhattan plots and quantile-quantile (QQ) plots from analysis of GWA for flushing with the relaxed definition in CKB.** Each plot represents the result from different models. (A) Model 1: controlling for age, age squared, PCs (1-10) (B) Model 2: covariates in model 1 plus ALDH2 rs671 and (C) Model 3: covariates in model 2 plus ADH1B rs1229984. The y axis shows the age and sex adjusted -log10 P values, and the x axis presents positions along the chromosome (Chr.). The solid red line indicates the P value of 5 x 10^-8^ where the blue line indicates the P value of 1 x 10^-5^. (D-F) represent QQ plots for each model.

**Supplementary Figure 2. Manhattan plots and quantile-quantile (QQ) plots from analysis of GWA for flushing with the strict definition in CKB.** Each plot represents the result from different models. (A) Model 1: controlling for age, age squared, PCs (1-10) (B) Model 2: covariates in model 1 plus ALDH2 rs671 and (C) Model 3: covariates in model 2 plus ADH1B rs1229984. The y axis shows the age and sex adjusted -log10 P values, and the x axis presents positions along the chromosome (Chr.). The solid red line indicates the P value of 5 x 10^-8^ where the blue line indicates the P value of 1 x 10^-5^. (D-F) represent QQ plots for each model.

**Supplementary Figure 3. Manhattan plots and quantile-quantile (QQ) plots from analysis of GWA for flushing with the continuous definition in CKB.** Each plot represents the result from different models. (A) Model 1: controlling for age, age squared, PCs (1-10) (B) Model 2: covariates in model 1 plus ALDH2 rs671 and (C) Model 3: covariates in model 2 plus ADH1B rs1229984. The y axis shows the age and sex adjusted -log10 P values, and the x axis presents positions along the chromosome (Chr.). The solid red line indicates the P value of 5 x 10^-8^ where the blue line indicates the P value of 1 x 10^-5^. (D-F) represent QQ plots for each model.

**Supplementary Figure 4. Manhattan plots and QQ plots of GWAS of alcohol flushing in KoGES.** Each plot represents the result from different models. (A and D) model 1: controlling for age, age squared, PCs (1-10) (B and E) model 2: covariates in model 1 plus ALDH2 rs671 and (C and F) model 3: covariates in model 2 plus ADH1B rs1229984. The y axis shows the age and sex adjusted -log10 P values, and the x axis presents positions along the chromosome (Chr.). The solid red line indicates the P value of 5 x 10^-8^ where the blue line indicates the P value of 1 x 10^-5^.

**Supplementary Figure 5. Manhattan plot of the results from the GWAS meta-analysis of alcohol flushing.** Each plot represents the result from different models. (A) Model 1: controlling for age, age squared, PCs (1-10) (B) Model 2: covariates in model 1 plus ALDH2 rs671 and (C) Model 3: covariates in model 2 plus ADH1B rs1229984. The y axis shows the age and sex adjusted -log10 P values, and the x axis presents positions along the chromosome (Chr.). The solid red line indicates the P value of 5 x 10^-8^ where the blue line indicates the P value of 1 x 10^-5^. (D-F) represent QQ plots for each model.

**Supplementary Figure 6. Regional association plot for the chromosome 12:110,000,000–114,000,000 region in CKB.** Plot show the most strongly associated SNP, rs671 (purple diamond). The plot is based on the meta-analysis result (model 1). The bottom panel indicates the genes and their orientation for each region.

**Supplementary Figure 7. A directed acyclic graph (DAG) of the model.** Alcohol intake is influenced by health status (e.g., higher levels of BP) and alcohol flushing. If the study subjects were selected by their drinking status, flushing and higher levels of BP are no longer independent, and biased estimates can be induced.

**Supplementary Figure 1. Manhattan plots and quantile-quantile (QQ) plots from analysis of GWA for flushing with the relaxed definition in CKB.**

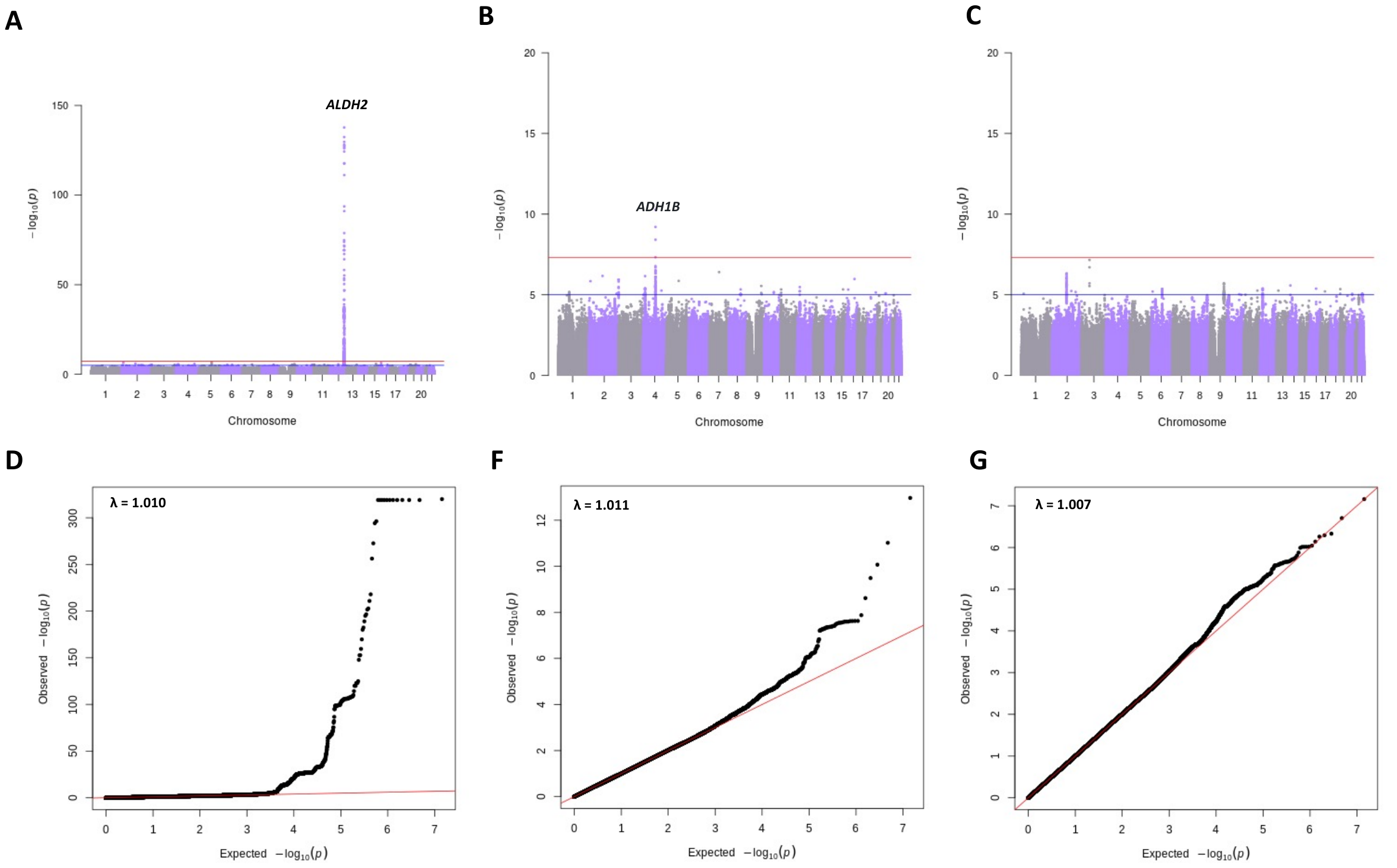

**Supplementary Figure 2. Manhattan plots and quantile-quantile (QQ) plots from analysis of GWA for flushing with the strict definition in CKB.**

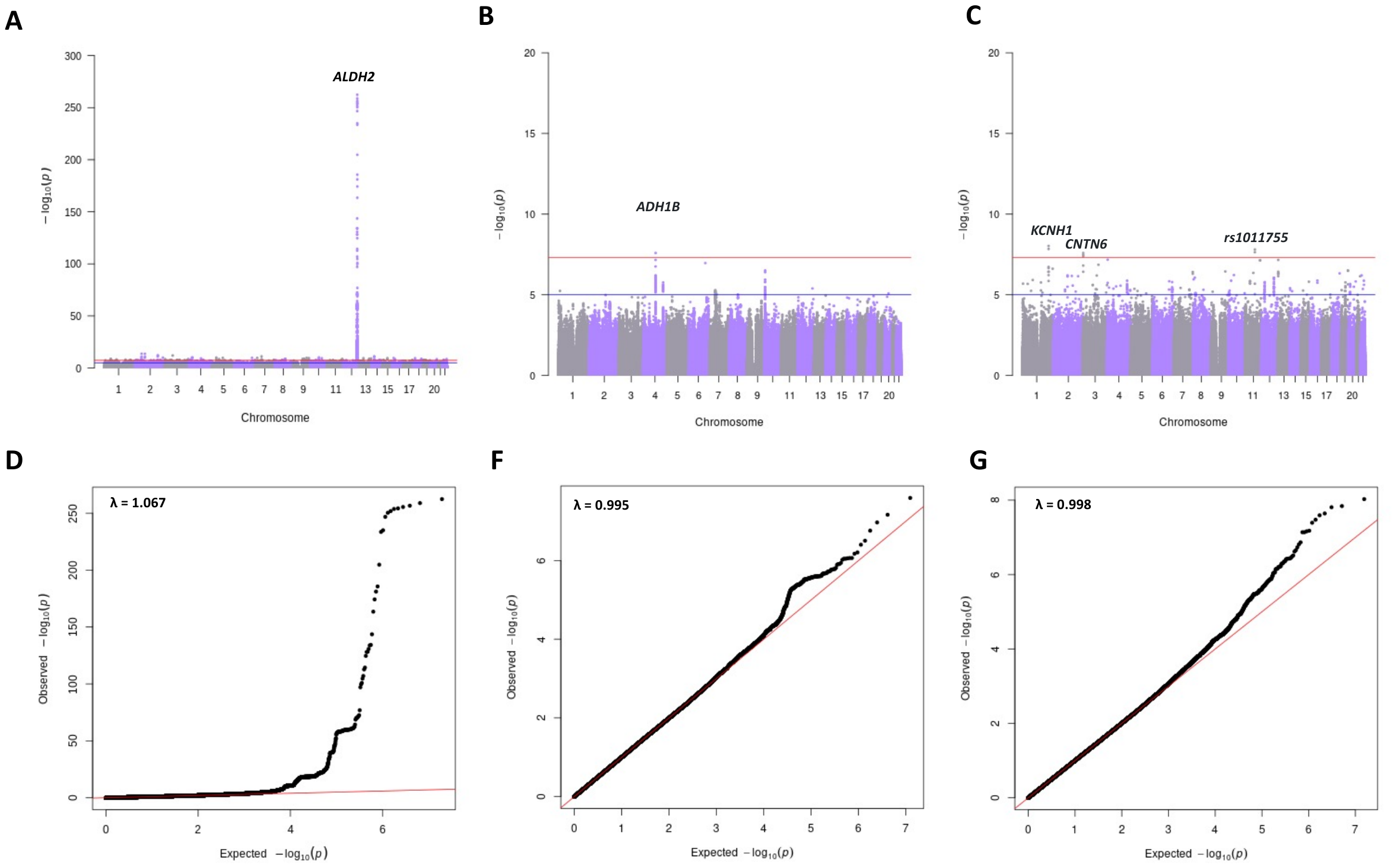

**Supplementary Figure 3. Manhattan plots and quantile-quantile (QQ) plots from analysis of GWA for flushing with the continuous definition in CKB.**

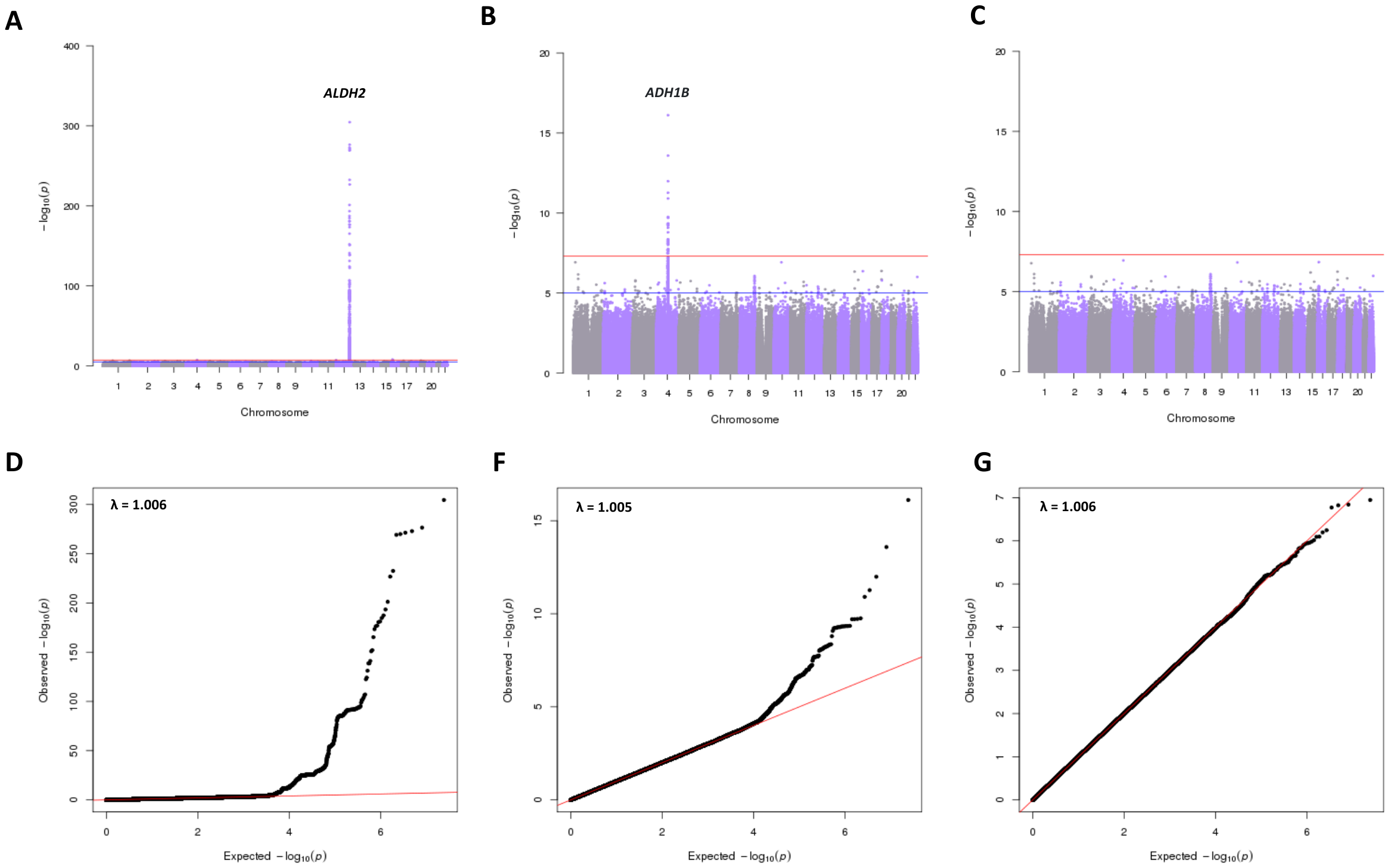

**Supplementary Figure 4. Manhattan plots and QQ plots of GWAS of alcohol flushing in KoGES.**

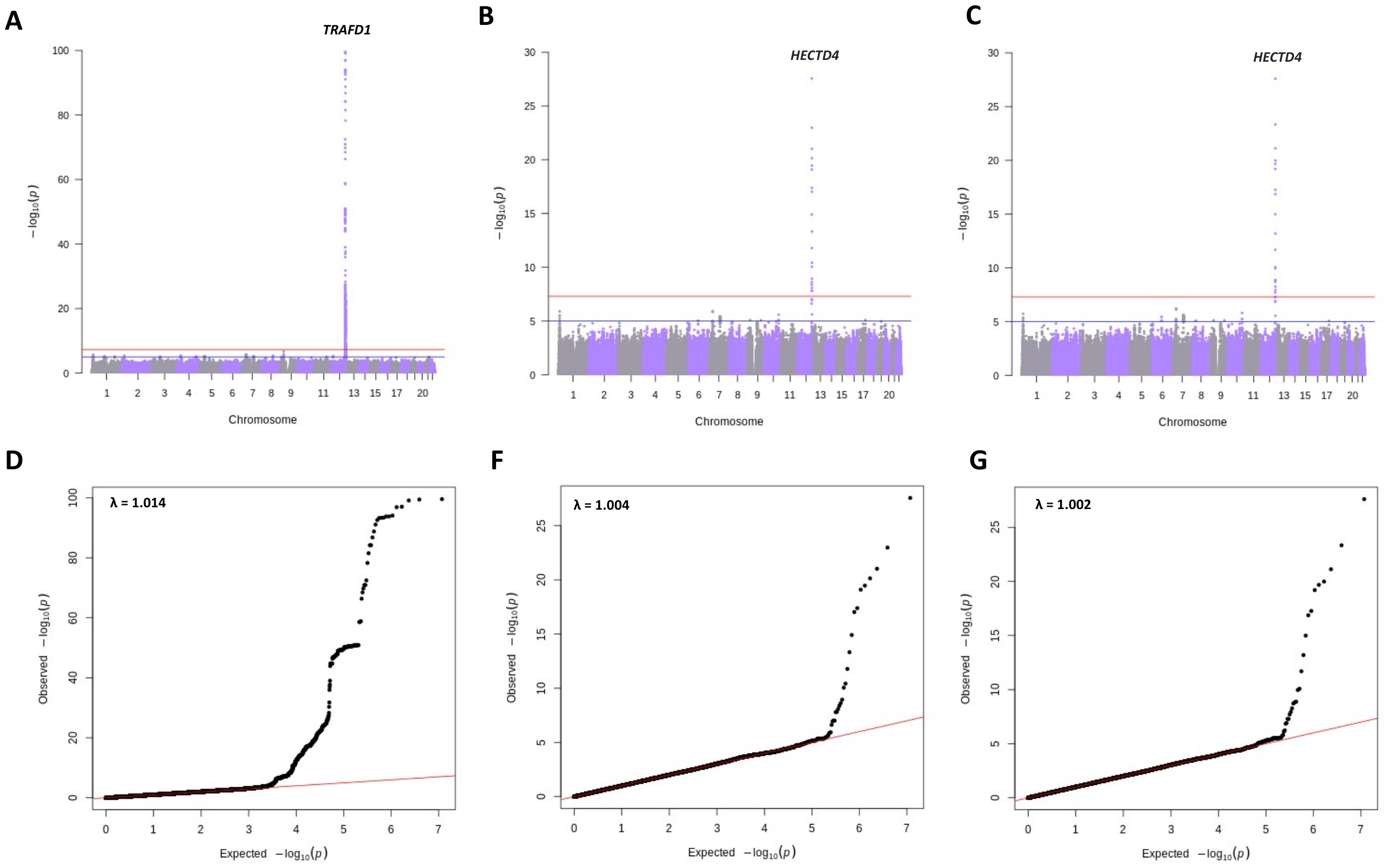

**Supplementary Figure 5. Manhattan plot of the results from the GWAS meta-analysis of alcohol flushing.**

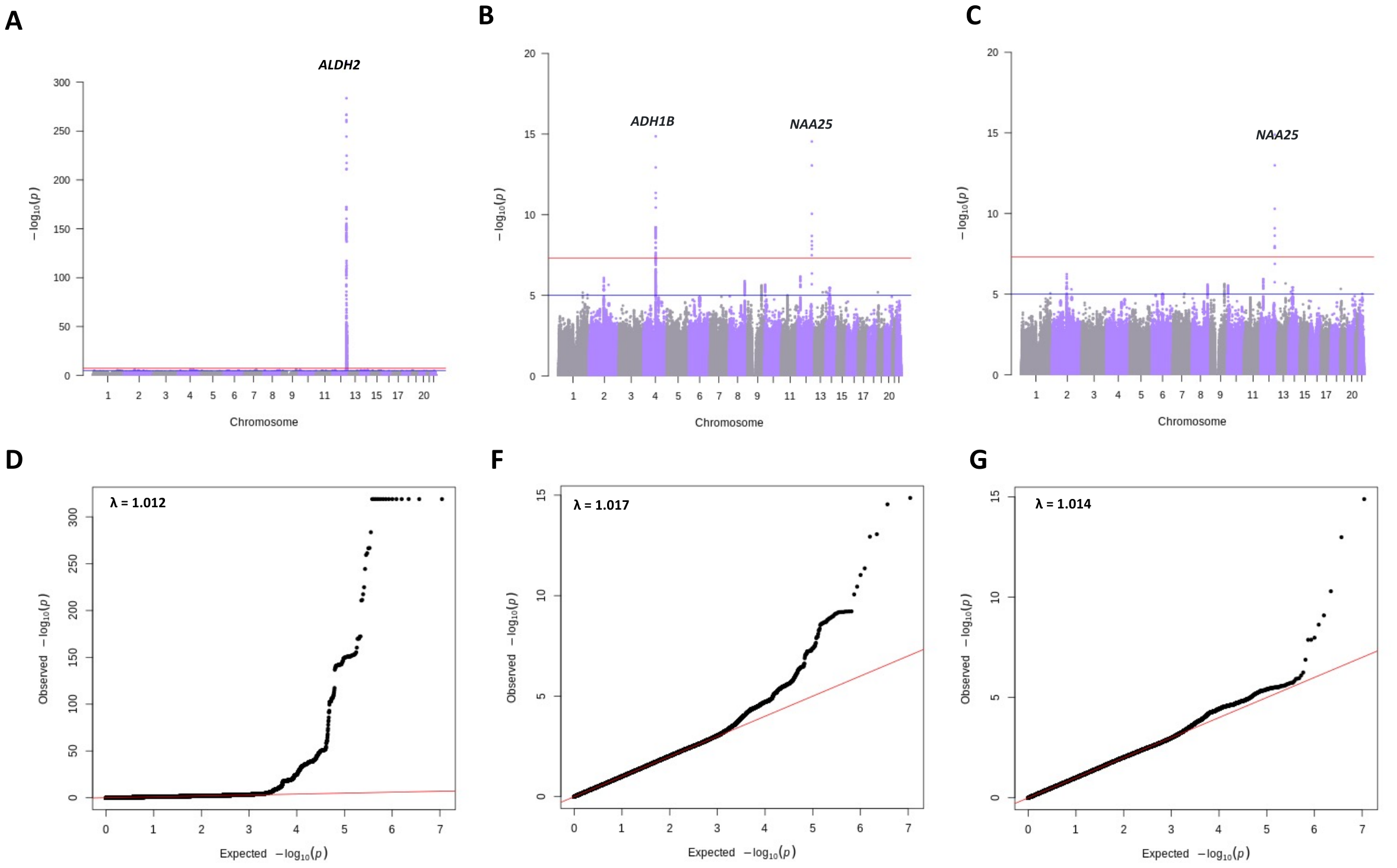

**Supplementary Figure 6. Regional association plot for the chromosome 12:110,000,000–114,000,000 region in CKB.**

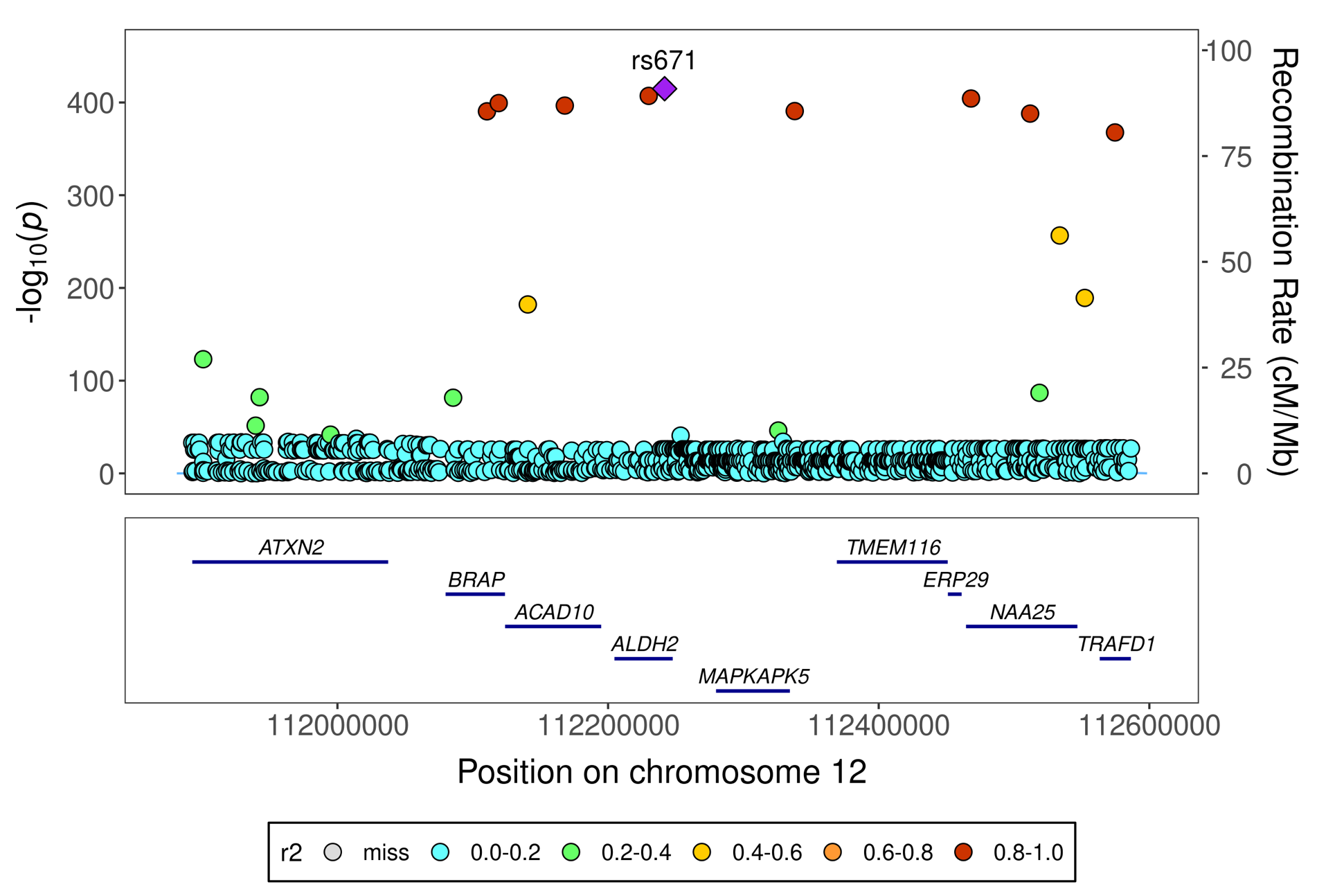

**Supplementary Figure 7. A directed acyclic graph (DAG) of the model.**

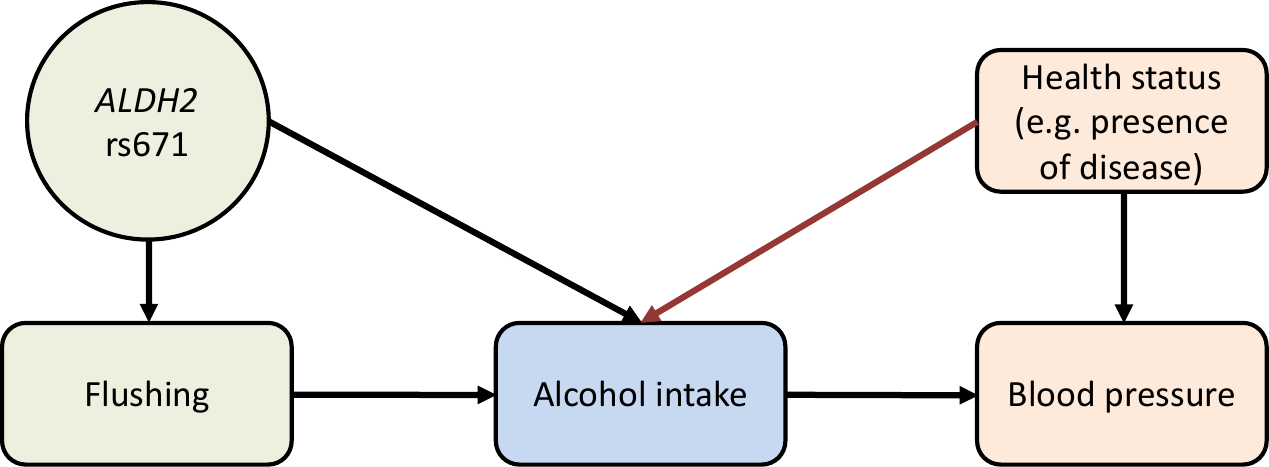

### **References**

1.	Kim, Y., B.G. Han, and G.E.S.g. Ko, *Cohort Profile: The Korean Genome and Epidemiology Study (KoGES) Consortium.* Int J Epidemiol, 2016.

2.	Baik, I. and C. Shin, *Prospective study of alcohol consumption and metabolic syndrome.* Am J Clin Nutr, 2008. **87**(5): p. 1455-63.

3.	Cho, Y.S., et al., *A large-scale genome-wide association study of Asian populations uncovers genetic factors influencing eight quantitative traits.* Nat Genet, 2009. **41**(5): p. 527-34.

4.	Genomes Project, C., et al., *A global reference for human genetic variation.* Nature, 2015. **526**(7571): p. 68-74.

5.	Yang, J., et al., *Common SNPs explain a large proportion of the heritability for human height.* Nat Genet, 2010. **42**(7): p. 565-9.

6.	Lee, S.H., et al., *Estimation of pleiotropy between complex diseases using single-nucleotide polymorphism-derived genomic relationships and restricted maximum likelihood.* Bioinformatics, 2012. **28**(19): p. 2540-2542.

7.	Wang, G., et al., *A simple new approach to variable selection in regression, with application to genetic fine mapping.* Journal of the Royal Statistical Society: Series B (Statistical Methodology), 2020. **82**(5): p. 1273-1300.
